# The neuronal chromatin landscape in adult schizophrenia brains is linked to early fetal development

**DOI:** 10.1101/2023.10.02.23296067

**Authors:** Kiran Girdhar, Jaroslav Bendl, Andrew Baumgartner, Karen Therrien, Sanan Venkatesh, Deepika Mathur, Pengfei Dong, Samir Rahman, Steven P. Kleopoulos, Ruth Misir, Sarah M. Reach, Pavan K. Auluck, Stefano Marenco, David A. Lewis, Vahram Haroutunian, Cory Funk, Georgios Voloudakis, Gabriel E. Hoffman, John F. Fullard, Panos Roussos

## Abstract

Non-coding variants increase risk of neuropsychiatric disease. However, our understanding of the cell-type specific role of the non-coding genome in disease is incomplete. We performed population scale (N=1,393) chromatin accessibility profiling of neurons and non-neurons from two neocortical brain regions: the anterior cingulate cortex and dorsolateral prefrontal cortex. Across both regions, we observed notable differences in neuronal chromatin accessibility between schizophrenia cases and controls. A per-sample disease pseudotime was positively associated with genetic liability for schizophrenia. Organizing chromatin into *cis*- and *trans*-regulatory domains, identified a prominent neuronal *trans*-regulatory domain (TRD1) active in immature glutamatergic neurons during fetal development. Polygenic risk score analysis using genetic variants within chromatin accessibility of TRD1 successfully predicted susceptibility to schizophrenia in the Million Veteran Program cohort. Overall, we present the most extensive resource to date of chromatin accessibility in the human cortex, yielding insights into the cell-type specific etiology of schizophrenia.

## Introduction

The epigenome encompasses the architecture of DNA packaged inside the nucleus, including chromatin accessibility, DNA methylation and a range of histone marks. This orchestration dynamically regulates gene expression throughout life. Marked by rapid and intricate changes, the early stages of brain development offer a critical window where genetic and environmental factors converge, ultimately influencing neural circuitry and pathways that shape behavior and cognition (*1–3*). The interplay between genetics and environment (GxE), plays also a pivotal role in neurodevelopmental disorders, including schizophrenia (*4*). Within this highly complex landscape, the epigenome emerges as a landmark for navigating the link between genetics, environment, and schizophrenia. Epigenetic marks not only reflect the combined influences of GxE, but also hold the potential to be modified by therapeutic interventions, offering new avenues for treatment strategies.

Investigation of the epigenetic architecture of schizophrenia using high-throughput assays such as the assay for transposase-accessible chromatin followed by sequencing (ATAC-seq) and chromatin immunoprecipitation sequencing (ChIP-seq) has been limited to two main studies. In the first study, ATAC-seq was applied in the homogenate prefrontal cortex (PFC) of 135 schizophrenia cases and 137 controls (*5*). In the second study, ChIP-seq for H3K27ac and H3K4me3 was performed on PFC neurons from more than ∼120 schizophrenia cases and a similar number of matched controls (*6*). Despite these valuable resources, these studies have certain limitations. Firstly, the representation of cell- and region-specific open chromatin regions (OCRs) is absent. Also, the disease specific chromatin landscape that encompasses all regulatory elements is incomplete as the markers used, H3K27ac and H3K4me3, only identify active enhancers and promoters.

We aimed to address these limitations by profiling, via ATAC-seq, cell- and brain-region-specific chromatin accessibility patterns in postmortem brain samples from 469 unique donors, comprising controls and individuals diagnosed with schizophrenia and bipolar disorder (BD). Fluorescence Activated Nuclei Sorting (FANS) was used to isolate neuronal and non-neuronal nuclei from two disease relevant brain regions: the PFC and the anterior cingulate cortex (ACC), creating the largest collection, to-date, of disease associated chromatin accessibility profiles in the human brain. We used this dataset to examine specific patterns of chromatin organization in different disease contexts. We focused on schizophrenia and interrogated the higher order chromatin structure spanned by *cis*- and *trans*-regulatory domains (CRDs and TRDs, respectively). Integration with chromatin accessibility from the human fetal cortex annotated the spatiotemporal patterns of schizophrenia-associated OCRs in TRDs. Finally, stratification of polygenic risk scores (PRS) using genetic variants within TRDs identified the functional sub-structure of higher order chromatin that predicts schizophrenia cases in an independent cohort from the Million Veteran Program. Due to the absence of strong findings associated with BD, we limited our downstream analysis to schizophrenia and controls.

## Results

### Population-scale chromatin accessibility data captures cell type-specific enhancer-promoter interactions in human cortical brain regions

To study the relevance of non-coding genomic regions to serious mental illness, we performed ATAC-seq on neuronal and non-neuronal nuclei in frozen postmortem tissues from PFC and ACC in a cohort of schizophrenia (n=157), BD (n=77) and controls (n=235) (**Fig. 1A, Fig. S1** and **Table S1A-E**). After quality control measures, we obtained a total of 1,393 libraries, collectively consisting of over 54.8 billion unique reads (**Fig. S2-S3, Table S1F,** and **methods**). Given that the majority of variance in OCR expression was attributed to cell type (**Fig. S4**) and considering the substantial differences in cell-type specific OCRs (**Fig. 1B**), along with significant overlap across cortical region specific OCRs, we examined neuronal and non-neuronal samples separately in subsequent downstream analysis.

**Figure 1:**
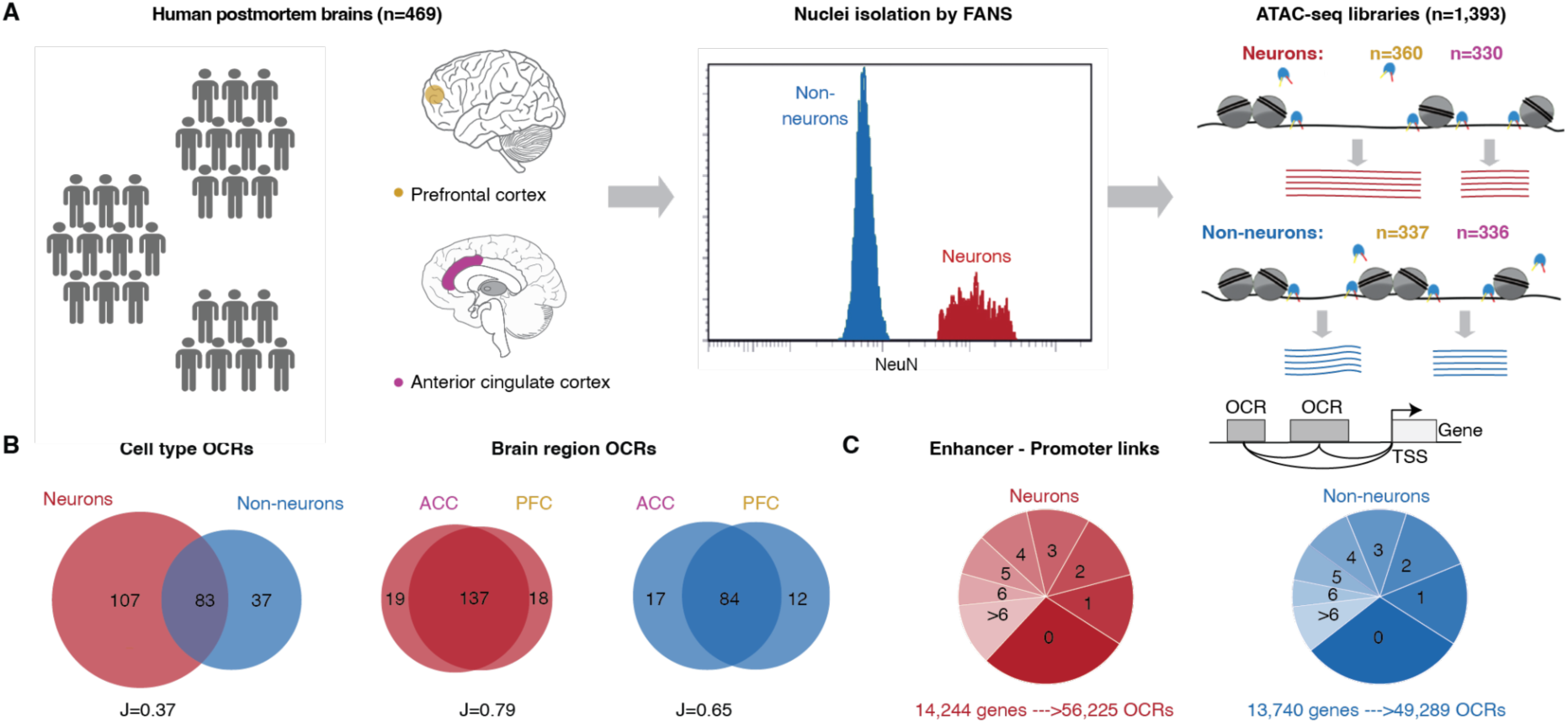
Population-scale chromatin accessibility analysis in the human brain. **A**) Brain tissue specimens were obtained from 469 unique donors, comprising individuals with schizophrenia (SCZ) (n=157), BD (n=77) and controls (n=235). Neuronal and non-neuronal nuclei were isolated by FANS and ATAC-seq profiling was performed to generate a total of 1,393 libraries. **B**) Venn diagram showing cell-type (left) and brain region (right) specificity of identified OCRs. **C**) Top: Schematic to show enhancer-promoter links. Grey box, light grey box and black arrow represent OCRs, TSS and gene body, respectively. Bottom: the distribution in pie charts show stratification of 19,749 genes annotated to the number of neuronal (shades of red) and non-neuronal (shades of blue) OCRs.

A total of 391,420 neuronal and 260,431 non-neuronal OCRs were identified, of which, the majority were distributed outside the transcription start site (TSS) (**Fig. S5A,** see **data and materials availability**). Over 83% of the OCRs identified in this study overlapped with previously observed regulatory elements from reference studies (*5*, *7–12*) (**Fig. S5D**), and the overlap was higher for matched cell types (**Fig. S5E**) and known cell type markers (**Fig. S5B**). The magnitude of chromatin accessibility was strongly correlated with an external ATAC-seq dataset from homogenate PFC tissue (*5*) (**Fig. S5C**).

Integration of cell specific OCRs with cell-type-matched ChIP-seq (*13*) and Hi-C (*12*) data, identified 56,225 neuron and 49,289 non-neuron enhancer-promoter (E-P) interactions using Activity-By-Contact (ABC) model (*14*), which linked distal OCRs, covering 17.7-18.6Mb in each cell type (0.60-0.64% of the genome), to 83% of the expressed genes (16,504 of 19,749) (**Fig. 1C and see data and materials availability** and **Fig. S6A**). While the majority of distal OCRs were predicted to interact with a single gene, and the frequency of E-P links decreased sharply with distance, about 39% of them were associated with two or more genes (**Fig. S6B, Fig. S6D**) and only about a quarter were linked to the nearest gene (**Fig. S6C**).

### Upregulated schizophrenia OCRs in neurons are enriched for schizophrenia risk variants

We next investigated changes in chromatin accessibility patterns associated with schizophrenia and BD separately for each brain region and cell-type, after correcting for covariates (**see methods, Fig. S7-8**). Notably, the highest number of schizophrenia-specific alterations (termed “schizophrenia OCRs”) were found in neurons of the PFC (41,387 OCRS, 10.5% of total), followed by the ACC (27,771/7.1% OCRs) (**Fig. 2A, Fig. S9A,** see **data and materials availability**). In contrast, non-neurons had significantly lower numbers of schizophrenia-specific alterations in both brain regions. The schizophrenia OCR changes in ACC and PFC were highly concordant (**Fig. S9B**), indicating comparable disease-associated epigenomic alterations in these regions. For BD, due to less power in the sample size, we found much lower numbers of associations (11-166 OCRs; under 0.1% OCRs) with small effect sizes (**Fig. S9A,** see **data and materials availability**).

**Figure 2:**
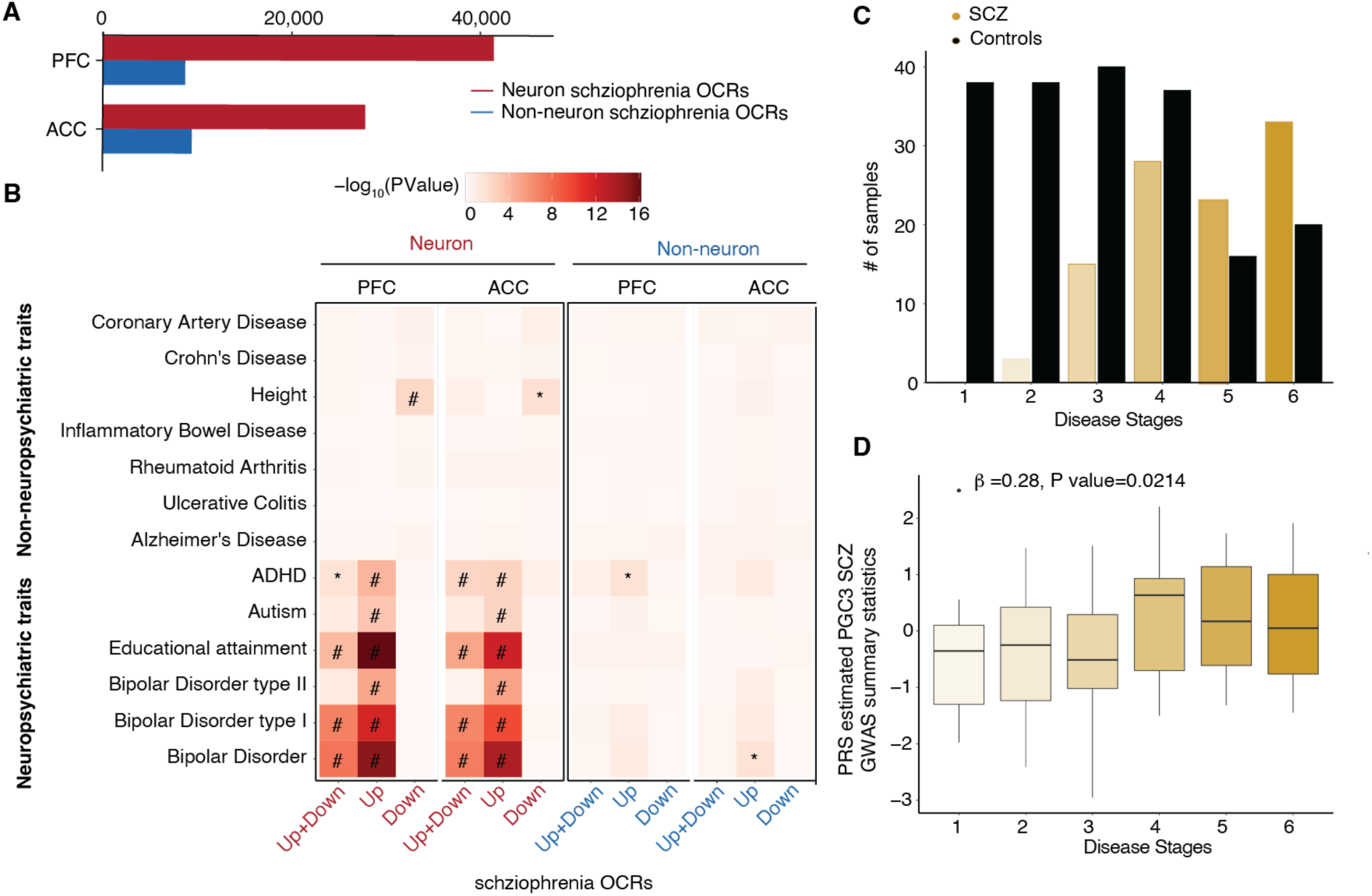
Schizophrenia OCRs and inferred schizophrenia stages in PFC neurons. **A**) Count of schizophrenia OCRs at FDR < .05 using differential analysis of schizophrenia vs controls for PFC and ACC in neurons and non-neurons. **B)** Heatmap of enrichment *P* values of neuropsychiatric and non-neuropsychiatric related GWAS traits. The overlap of OCRs with genetic variants was assessed using LD score regression. ‘#’: significant for enrichment in LD score regression after FDR correction of multiple testing across all tests in the plot (Benjamini–Hochberg test); ‘*’: nominally significant for enrichment. The heritability coefficients of common risk variants that overlap with (1) “Up+Down”: upregulated and downregulated schizophrenia OCRs (41,387(27,771) in PFC(ACC) neurons (red), 8,671(9,405) in PFC(ACC) non-neurons (blue)) (2) “Up”: upregulated schizophrenia OCRs (25,146(15,791) in PFC(ACC) in neurons (red), 3,726(3,075) in PFC(ACC) non-neurons (blue)); and (3) “Down”: dysregulated schizophrenia OCRs (16,241(11,980) in PFC(ACC) neurons (red), 4,945(6,330) in PFC(ACC) non-neurons (blue)). All upregulated (Up) and downregulated (Down) OCRs are with log2FC (schizophrenia versus controls) >0 and <0, respectively. **C**) Stratification of neuronal PFC samples by clinical diagnosis as a function of inferred disease stage, where early stage (n=1) to late stage (n=6) is from left to right. **D**) Plot of PRS of neuronal PFC samples calculated using Psychiatric Genomics Consortium (PGC3) schizophrenia GWAS summary statistics, stratified by inferred disease severity stages in neurons from PFC region. Beta estimate and p value are obtained using linear regression model: disease stages ∼ PRS + Age + Age^2^ + Sex. Box plots have lower and upper hinges at the 25th and 75th percentiles and whiskers extending to, at most, 1.5xIQR (interquartile range).

We then tested whether the observed schizophrenia OCRs were enriched for schizophrenia risk variants. We found magnitude of schizophrenia heritability in neurons was almost two fold for differentially upregulated OCRs in PFC and ACC than all schizophrenia OCRs, whereas downregulated OCRs in neurons had negative and insignificant coefficients in both regions (**Fig. S10A, Fig. 2B**). The co-localization of schizophrenia risk variants, and other co-heritable traits, such as BD type I, educational attainment and attention deficit hyperactivity disorder (ADHD) with neuron-specific OCRs is in line with the well-established fact that neuropsychiatric traits are enriched in neuronal, rather than non-neuronal, cell populations (*16–18*) (*19*) and also exhibited enrichment in upregulated schizophrenia OCRs (**Fig. 2B**, **Table S2**).

### Severity of schizophrenia inferred from expression of disease open chromatin regions

Having shown that upregulated schizophrenia OCRs in neurons are enriched with schizophrenia risk variants, we next used the manifold learning method previously employed to investigate Alzheimer’s disease progression in bulk RNA-seq data from postmortem brains (*20*) to infer disease pseudotime or stages, for each sample. After obtaining the nonlinear function of OCR expression embedded in low-dimensional space, the samples were ordered based on the similarity in expression of upregulated schizophrenia OCRs. Leiden clustering was applied to stratify the samples into six(five) distinct groups in neurons in PFC(ACC) regions denoting an ascending progression of disease severity as shown in **Fig. S11A(B) (**see **methods**). This is depicted by control samples being better aligned with less severe disease stages whereas schizophrenia cases were disproportionately represented in groups associated with more severe disease stages in PFC (**Fig. 2C)** and ACC (**Fig. S11C**) (see **Table S3A-B**).

To evaluate the accuracy of the inferred disease stages as a model of disease progression, we calculated a metric called diffusion pseudotime (*20*, *21*) (see **methods**). We leveraged the pseudotime metric to measure the severity of disease for two groups (schizophrenia and controls) for each brain cortical region. Interestingly, the odds of observing schizophrenia individuals at later pseudotime values from the disease trajectory of neurons is greater than one when compared to controls who have mainly early pseudotime values across both cortical regions (**Fig. S11D-E**), while no association was found between the inferred disease stages and other demographic and technical measures of samples (**Fig. S12**).

To measure the link between genetic factors and disease stages, we first estimated schizophrenia PRS for all samples and estimated the correlation with disease status across PFC and ACC region. **Figure S11F-G** show positive and significant association of PRS with disease status in our study. Next, we hypothesized that per sample inferred disease severity is partially explained by schizophrenia PRS. We observed a significant positive correlation between schizophrenia PRS and disease stages in the PFC (β=0.28, P value = 0.0214, n=140) (**Fig. 2D** and **Table S3C**) and ACC ((β=0.27, P value = 0.0172, n=132) using the model disease stages ∼ PRS + Age + Age^2^ + Sex (**Fig. S11H** and **Table S3D**), providing a convergence of the degree of perturbation in the epigenome of schizophrenia cases with disease heritability.

### Open chromatin regions in *cis*-regulatory landscape colocalize with schizophrenia risk loci

Genomic regulatory elements physically interact at multiple resolutions within the three-dimensional genome for precise gene regulation and overall functioning of cells. Here we focus on identifying physically interacting regulatory elements, named *cis*-regulatory domains (CRDs) which are shown to be delimited by topologically associating domain (TAD) and subTAD boundaries and enriched in binding sites for CCCTC binding factor (CTCF). These domains can be estimated using inter-individual variations in E-P associated histone marks and cell-type specific OCRs (*6*, *22–24*). Using the same analytical framework (see **methods**, **Fig. S13**), ∼37% of neuronal OCRs were assembled into 6,706 PFC and 6,625 ACC CRDs, and ∼33% of non-neuronal OCRs assembled into 4,612 PFC and 4,710 ACC CRDs (**Fig. S14A**). On average, 21.7(22.1) and 18.9(18.7) OCRs per CRD were present in neuronal and non-neuronal CRDs in PFC(ACC) respectively (**Fig. S15A** and **Table S4**). OCRs within CRDs showed cell and region specificity (**Fig. S15B**). Neuronal and non-neuronal CRDs had a median length of 27.7Kb(26.9Kb) and 42.7Kb(40.3Kb) in PFC(ACC), respectively (**Fig. S14B**). The identified CRDs were more likely to be located inside cell specific subTADs (**Fig. S15C**), and CRD borders exhibited strong enrichment for CTCF binding sites (**Fig. S15D-H**). E-P interactions showed significant enrichment in neuronal and non-neuronal CRDs compared to all OCRs **(Fig. S15I**). Taken together, cell and region specific CRDs are small structural subunits that constitute E-P interactions which are delimited by subTADs and exhibit characteristics of three dimensional genome organization.

We examined schizophrenia heritability for OCRs that are inside and outside CRDs. Interestingly, only OCRs inside neuronal CRDs show significant schizophrenia heritability (**Fig. 3A** and **Table S5**). Next, we compared the relationship among the disease-associated OCRs, CRDs and “schizophrenia CRDs”. We observed neuronal OCRs with disease signatures to be more likely inside than outside CRDs, based on disease associated t-stats of OCRs (OR =1.331(1.338), *P* < 2.2e-16(2.2e-16) in PFC(ACC)) using logistic regression based generalized linear model (GLM) (see **methods**). On the other hand, non-neuronal CRDs, showed depletion for disease associated OCRs (GLM results: OR =0.961(0.965), *P* = 1.62e-58(3.07e-45) in PFC(ACC)) (**Fig. S16A-B**). We then compared the disease-associated OCRs with identified 1,953(1,691) neuronal and 1,003(1,090) non-neuronal “schizophrenia CRDs” that exhibit dysregulation in their mean expression between individuals with schizophrenia and controls in PFC(ACC) (**Fig. 3B and methods,** see **data and materials availability**). Genome-wide, approximately 44.9%(41.9%), corresponding to 18,598(11,663) out of 41,387(27,771) neuronal schizophrenia OCRs were found to be encompassed within neuronal PFC(ACC) derived schizophrenia associated CRDs. Similarly, approximately 30.6%(27.3%) of the non-neuronal schizophrenia OCRs, amounting to 2,656(2,569) out of 8,671(9,405), were identified as being contained within non-neuronal schizophrenia CRDs. Across all four datasets, schizophrenia OCRs tend to be clustered inside schizophrenia CRDs rather than outside schizophrenia CRDs (**Fig. S16C**). Within the neuronal schizophrenia CRDs, a higher proportion of schizophrenia OCRs is linked to disease associated transcripts (**Fig. S16D-E**). Schizophrenia OCRs in neuronal schizophrenia CRDs had over 1.5-fold increase in schizophrenia heritability compared to all schizophrenia OCRs across the PFC(ACC) regions (**Fig. 3C)**, while schizophrenia OCRs outside neuronal schizophrenia CRDs had no significant schizophrenia heritability **(Table S6**). These findings suggest chromatin interactions from 37% of OCRs that comprise neuronal CRDs delimit the OCRs that are significantly enriched for schizophrenia risk loci from the rest of the genome, and that these can be explored to identify disease associated expression changes.

**Figure 3:**
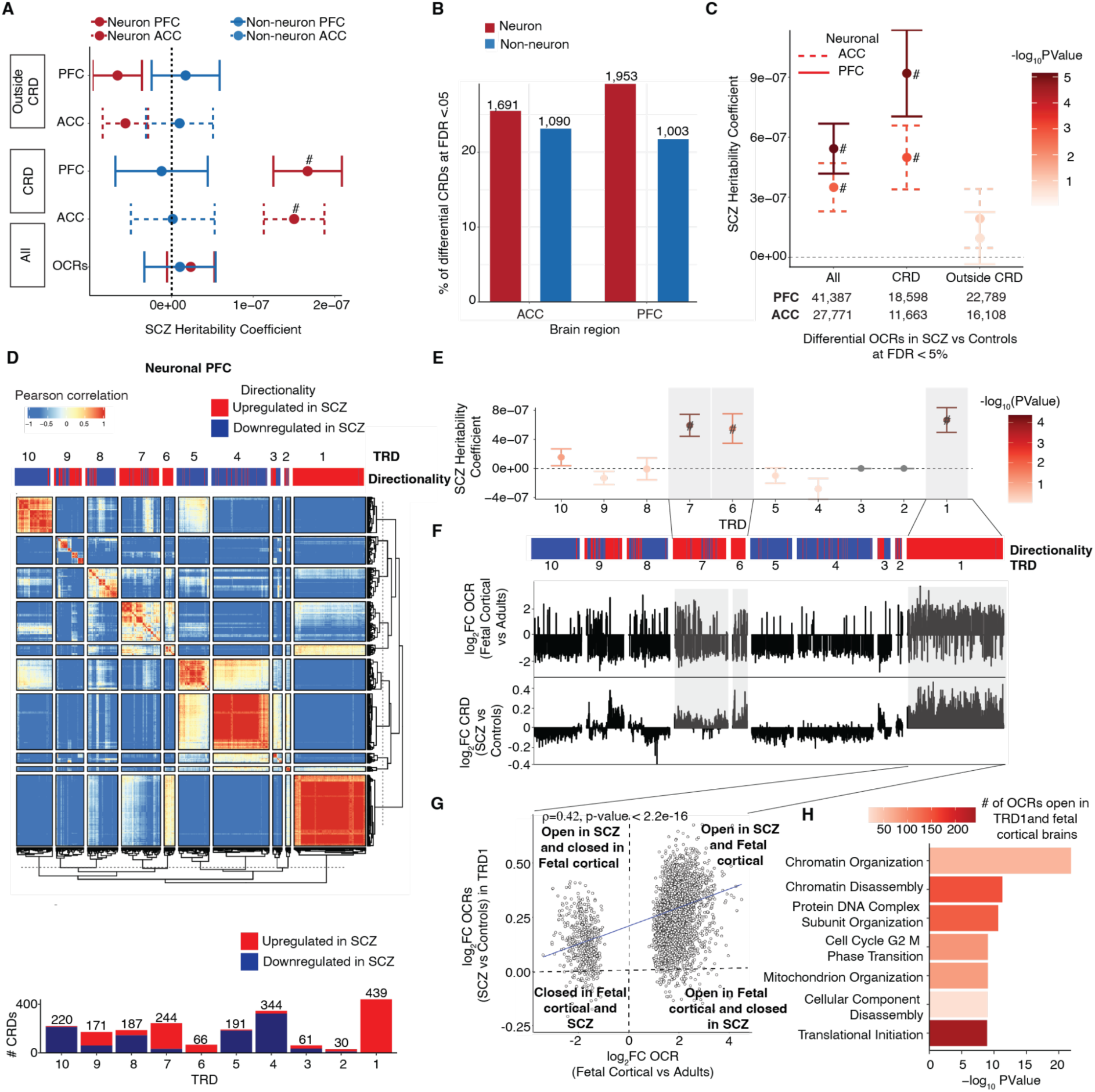
*Cis*-regulatory landscape and trans regulatory domains from hierarchical clustering of neuronal schizophrenia CRDs from the PFC. **A)** Schizophrenia heritability coefficients for neurons (red) and for non-neurons (blue) stratified by (1) “All”: all OCRs (neuronal = 391,420 and non-neuronal =260,431), (2) “CRD”: OCRs inside CRDs (neuronal = 145,533(146,148) and non-neuronal 87,222(87918) in PFC(ACC)) (3) “Outside CRD”: OCRs outside CRDs (neuronal = 245,272(245,887) and non-neuronal 173,209(172,513) in PFC(ACC)). **B**) Bar plot of number of differential CRDs across four datasets identified using a two-stage test at FDR<5%. **C**) Schizophrenia heritability coefficients for neurons (red) and non-neurons (blue) stratified by (1) “All”: all schizophrenia OCRs (2) “CRD”: schizophrenia OCRs inside schizophrenia CRDs and (3) “Outside CRDs”: schizophrenia OCRs outside schizophrenia CRDs. schizophrenia OCRs are identified from the previous section (see Fig. 2) at FDR 5%. **D**) Heatmap depicting hierarchical clustering of trans interactions of 1,953 schizophrenia CRDs across 360 samples from the PFC. The clustering results in ten trans regulatory domains (TRDs) using gamma statistics. The directionality annotation bar plot above the heatmap shows upregulation in red (log2SCZ-log2Control >0) and downregulation in navy (log2SCZ-log2Control <0) of schizophrenia neuronal CRDs. Bar plot of Number of up and downregulated schizophrenia neuronal CRDs per TRD from the PFC region. **E**) Coefficients of schizophrenia heritability stratified by TRDs. **F**) Top and bottom bar plots show log2FCfetal_cortical (Fetal cortical vs adult controls) and log2FC (schizophrenia vs Controls) of neuronal PFC CRD respectively. **G**) Spearman correlation of log2FCfetal_cortical compared to log2FC (schizophrenia vs Controls) of neuronal PFC CRDs from TRD1 (n=3,056 OCRs). P value is obtained from Spearman’s rank correlation (ρ) test. **H**) Functional pathway enrichment of OCRs from neuronal PFC TRD1 that overlap with fetal cortical specific OCRs. The overlap of OCRs with schizophrenia risk variants A), C) and E) were assessed using LD score regression. *P* values are from LD score regression. ‘#’: significant for enrichment in LD score regression after Benjamini–Hochberg FDR correction for multiple testing across all tests in the plot (FDR < 5%). Error bars show standard error in schizophrenia heritability from LD score regression.

### Trans interactions of CRDs in schizophrenia brains are enriched for neurodevelopmental OCRs

We next examined whether the interaction between disease associated CRDs could identify trans-regulatory domains (TRDs) that act as hubs for convergence among the degree of perturbation in the epigenome of schizophrenia cases with disease heritability. The analytical workflow from CRD to TRDs is shown in schematic displayed in **Fig. S17A**. We applied hierarchical clustering on a correlation matrix of expression of schizophrenia CRDs across all neuronal samples and Gamma statistics (*25*) to identify a total of 10 neuronal TRDs (**see methods** and **Fig. S17B**). While most TRDs exhibit a mix of upregulated and downregulated CRDs, there is one particular TRD (TRD1) that stands out with a predominant upregulation of CRDs in PFC and ACC (**Fig. 3D** and **Fig. S18A;** see **data and materials availability**). TRD1 also had the highest proportion of schizophrenia OCRs compared to other TRDs (**Fig. S17C**). We then examined the association of TRDs with schizophrenia risk loci. Three TRDs (1, 6, 7) out of ten in PFC (**Fig. 3E and see data and materials availability**) and two TRDs (1, 2) out of ten in ACC (**Fig. S18B** and see **data and materials availability**) had significant and positive coefficients of schizophrenia heritability for OCRs contained within them. One of the common characteristics of TRDs (1, 6, 7) in PFC and TRDs (1,2) in ACC, referred to as “schizophrenia TRDs” hereon, was a higher proportion of upregulated schizophrenia OCRs out of the total number of schizophrenia OCRs contained within them (**Fig. S17D**). To connect these OCRs with molecular mechanisms, we performed functional pathway analysis of OCRs in “schizophrenia TRDs”. Across PFC and ACC, TRD1 shows enrichment in biological processes such as membrane organization, cellular component organization, synaptic maturation and regulation of synaptic plasticity (see **data and materials availability**). PFC TRDs 6 and 7, and ACC TRD2 showed enrichment for the neuronal system, Rho GTPases and signalling proteins. In summary, we noticed multiple neurodevelopmental signatures such as chromatin organization and synaptic maturation in TRDs, however, it wasn’t clear which specific molecular mechanism was present in a specific TRD.

Given the enrichment of schizophrenia TRDs with neurodevelopmental signatures such as chromatin organization and synaptic maturation, we hypothesized that schizophrenia TRDs contain OCRs that are active during neurodevelopment. To explore this hypothesis, we first performed ATAC-seq in fetal cortical plate (post-conception weeks: 19-24; n=4 samples) (**see methods**) and compared it with adult controls from PFC neurons (aged >30 years, n=6) (29) to identify OCRs specific to early brain development. Upon comparison with fetal cortical samples, the upregulation in schizophrenia associated changes in CRDs within TRDs was strongly correlated with upregulation in early brain development OCRs in the PFC (**Fig. 3F**) and ACC (**Fig. S18C**). Thus, chromatin regions that are open in fetal cortical samples, are more likely to also be open in adult schizophrenia samples than in adult controls. Comparison of development-specific OCRs with schizophrenia associated changes in OCRs showed a significant correlation in TRD1 (ρ=0.42 at p value < 2.2e-16, n=3,056 OCRs) in PFC (**Fig. 3G**) and (ρ=0.46 at p value < 2.2e-16, n=3,149 OCRs) in ACC (**Fig. S18D**). In the PFC, the correlation of TRD6 with developmental-specific OCRs was slightly weaker (ρ=0.24, p value=3.47e-06, n=371 OCRs). However, no significant correlation was observed in PFC-TRD7 (ρ=0.02, p value=0.587, n=523 OCRs) and ACC-TRD2 (ρ=0.11, p value=.04, n=371 OCRs). Functional pathway analysis in OCRs that are upregulated in fetal cortical samples and PFC-TRD1, ACC-TRD1 and PFC-TRD6 identified biological processes related to cell adhesion, translation initiation, chromatin organization and chromatin disassembly (**Fig. 3H, Fig. S18E,** and see **data and materials availability**).

### Schizophrenia-specific neurodevelopmental TRDs are associated with early and late stages of brain development

After observing the neurodevelopmental signatures in neuronal schizophrenia TRDs, we aimed to identify specific neuronal cell types and brain developmental stages where these disease-associated perturbations occur. To accomplish this, we tested whether OCRs in neuronal TRDs are expressed in fetal cell types using an atlas of single-nucleus ATAC-seq data generated from human fetal cortical samples spanning 8 weeks across mid-gestation: specifically, at pcw16, 20, 21, and 24 (see **methods**) (*26*). The early stages (pcw16 and 20) are characterized by cell division and neural precursor proliferation, whereas the later stages (pcw21 and 24) are associated with cell migration and maturation. Firstly, the schizophrenia OCRs in fetal cell types displayed positive z-scores for TRD1 in both PFC and ACC (**Fig. 4A** and **Fig. S19A**), compared to other schizophrenia TRDs (**Fig. S20**). Secondly, TRD1 consistently showed positive and significant differential enrichment scores in GluN(1,3,5,8) across PFC and ACC during early fetal stages of brain development (pcw16+20). In contrast, GluN(6,9) showed significant association at later stages of development (pcw21+24) (**Fig. 4B** and **Fig. S19B**). Regarding inhibitory neurons, IN(3,4,5) displayed positive and significant differential enrichment scores in PFC and ACC at all stages of brain development (**Fig. 4C** and **Fig. S19C**). None of the other schizophrenia TRDs showed association with fetal cell types (**Table S7A-B**).

**Figure 4:**
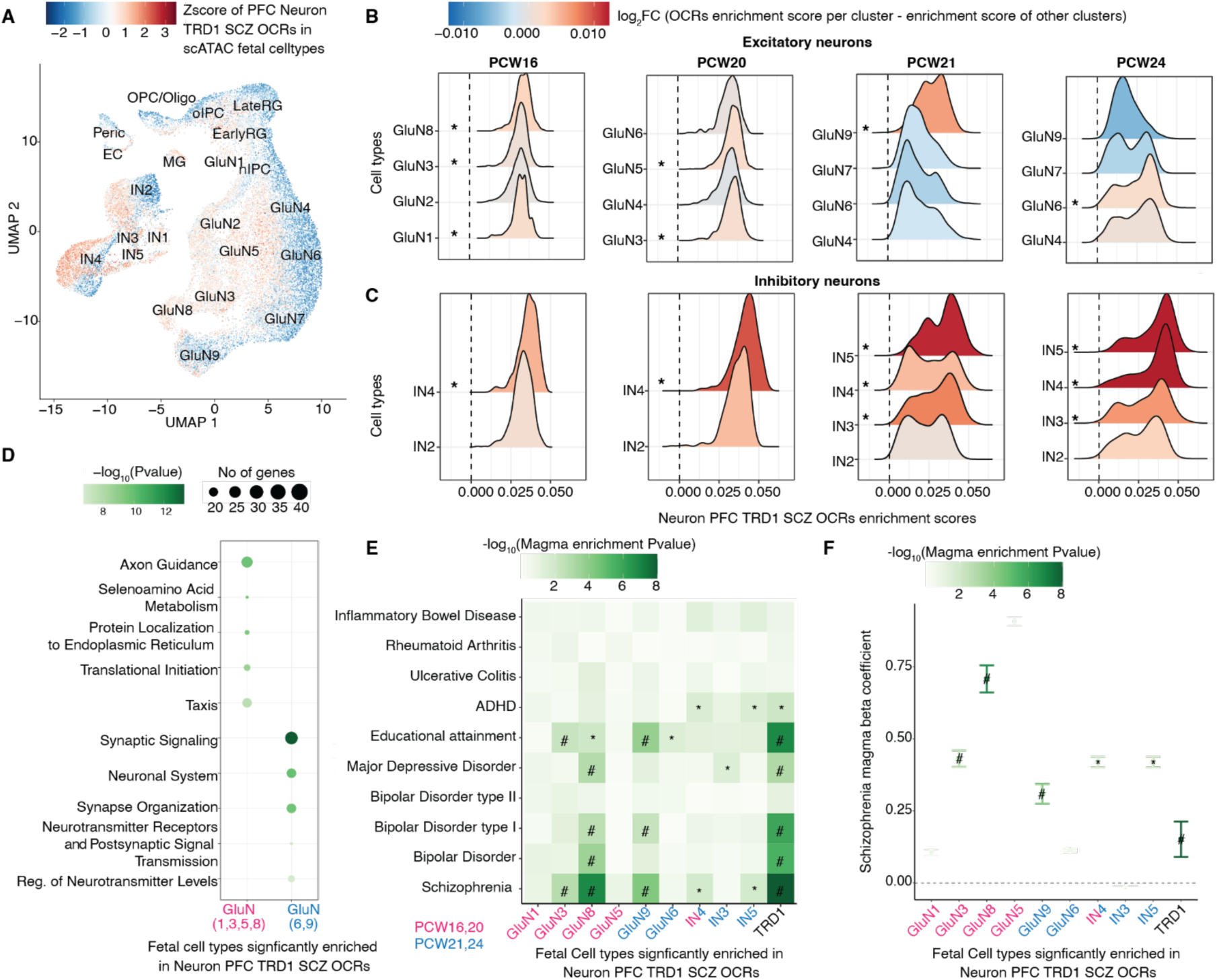
Analysis of cell-type specificity of neuronal PFC schizophrenia OCRs in fetal cortical scATAC-seq data. **A**) Uniform Manifold Approximation Projection (UMAP) plot of fetal cortical scATAC-seq data from Trevino *et al.* in which each cell type is colored by Z-scores of schizophrenia OCRs in neuronal PFC TRD1. The cell types include early radial glia (EarlyRG), late radial glia (LateRG), oligodendrocyte progenitor cell/oligodendrocyte (OPC/Oligo), neuronal intermediate progenitor cell (nIPC), oligo intermediate progenitor cell (oIPC), glutamatergic neuron (GluN), interneuron (IN), endothelial cell (EC), microglia (MG), and pericytes (Peric). Distribution of enrichment scores stratified by two major classes of cell types: **B**) excitatory neurons (nine subtypes of glutamatergic neurons) and **C**) inhibitory neurons (five subtypes of inhibitory neurons). The color represents the magnitude of coefficient of association of enrichment scores of schizophrenia OCRs from neuronal PFC-TRD1 at each developmental stage separately for each cell types, using the model (enrichment score ∼ celltype + (1|sample ID) where cell types are grouped as the cell type of interest vs. all other cell types. * depicts the cell types that are significantly enriched from the latter test and also have higher enrichment than the schizophrenia OCRs outside TRDs. **D**) Functional pathway analysis conducted on the early and late fetal glutamatergic specific marker genes that are also annotated by schizophrenia OCRs in TRD1. **E**) Heat map of MAGMA enrichment *P* values of brain-related and non-brain related GWAS traits and **F**) Coefficients of enrichment of common schizophrenia risk variants using MAGMA in fetal cell specific marker genes that are also annotated to schizophrenia OCRs within neuronal PFC TRD1. The x-axes in **E**) and **F**) are 1) early fetal (pcw16+20) glutamatergic (GluN 1,3,5,8) (magenta), 2) late fetal (pcw21+24) glutamatergic (GluN 6,9) (blue), 3) early+late fetal (pcw16+20+21+24) inhibitory (IN 4) (magenta+blue), 4) late fetal (pcw21+24) inhibitory (IN 3,5) fetal cell types (blue) and all ABC mapped schizophrenia OCRs to genes within neuronal PFC TRD1 (black). ‘#’: significant MAGMA enrichment coefficient after FDR correction of multiple testing across all tests in the plot (Benjamini–Hochberg test); ‘*’: nominally significant for enrichment in **E**) and **F**).

Next, we explored the specific molecular mechanism underlying these developmental stage specific glutamatergic, and inhibitory neurons, by intersecting cell types marker genes with E-P linked genes within TRD1 (see **methods**). Schizophrenia OCRs within TRD1 mapped to early stage genes from GluN(1,3,5,8) were enriched for DNA transcription and cell cycle processes (**Fig. 4D** and **Fig. S19D**); the late stages genes from GluN(6,9) were associated with synapse organization and maturation. On the other hand, schizophrenia OCRs within neuronal TRD1 mapped to early and late stage inhibitory neuronal genes were less in number resulting in underpowered pathway analysis that failed to pass multiple testing correction (**Table S7I-J**). Lastly, we asked whether these developmental cell specific genes have significant enrichment for schizophrenia risk loci. Remarkably, Glu (3,8,9) and IN(4,5) showed significant and positive coefficient of enrichment for schizophrenia risk loci and other co-heritable neurodevelopmental traits, including BD, ADHD, and educational attainment. No significant enrichment was observed for non-brain related traits, such as inflammatory bowel disease and rheumatoid arthritis. Taken together, these findings show that TRD1 isolates a set of OCRs that are upregulated in adult schizophrenia brains, are specific to neurodevelopmental glutamatergic and inhibitory neurons, and correspond to disrupted DNA binding related cellular processes during early stages and misregulated synaptic maturation during late stages of brain development.

### Neurodevelopmental TRDs stratify the schizophrenia cases and controls using polygenic risk scores

Finally, we explored whether the cumulative effect of genetic variation underlying schizophrenia TRD predicts an individual’s genetic susceptibility to schizophrenia and co-heritable traits, including BD and MDD in an independent cohort from the Million Veterans Program (MVP), which includes 1,957 (7,177; 120,067) schizophrenia (BD; MD) non-overlapping cases and 194,019 controls. Remarkably, PRS scores constructed with variants within schizophrenia OCRs within TRD1 (subsetted from the full PGC3 SCZ GWAS summary statistics) showed the highest odds of association with SCZ diagnosis when compared to other schizophrenia TRDs in the MVP cohort (**Fig. 5A-B**). After accounting for the varying number of genetic variants within each TRD (see **methods**), TRD1 showed the highest average variance explained per variant compared to other schizophrenia TRDs across both cortical regions and variants across the whole genome (**Fig. S21A**). We replicated this main finding in the CommonMind cohort (312 SCZ cases and 320 controls), where the variants withins OCRs in TRD1 showed the highest odds of classifying cases and controls and the highest average variance explained per variant in estimating PRS when compared to other schizophrenia TRDs (**Fig. S21B-C**). We observed a similarly strong correlation between PRS and stages in the PFC (Spearman rho=0.17, P value = 0.044, n=140) (**Fig. S21D**) and ACC (Spearman rho=0.21, P value = 0.013, n=132) (**Fig. S21E**), further pointing to the convergence of disease severity based on epigenetic dysregulation with genetic liability for schizophrenia.

**Figure 5:**
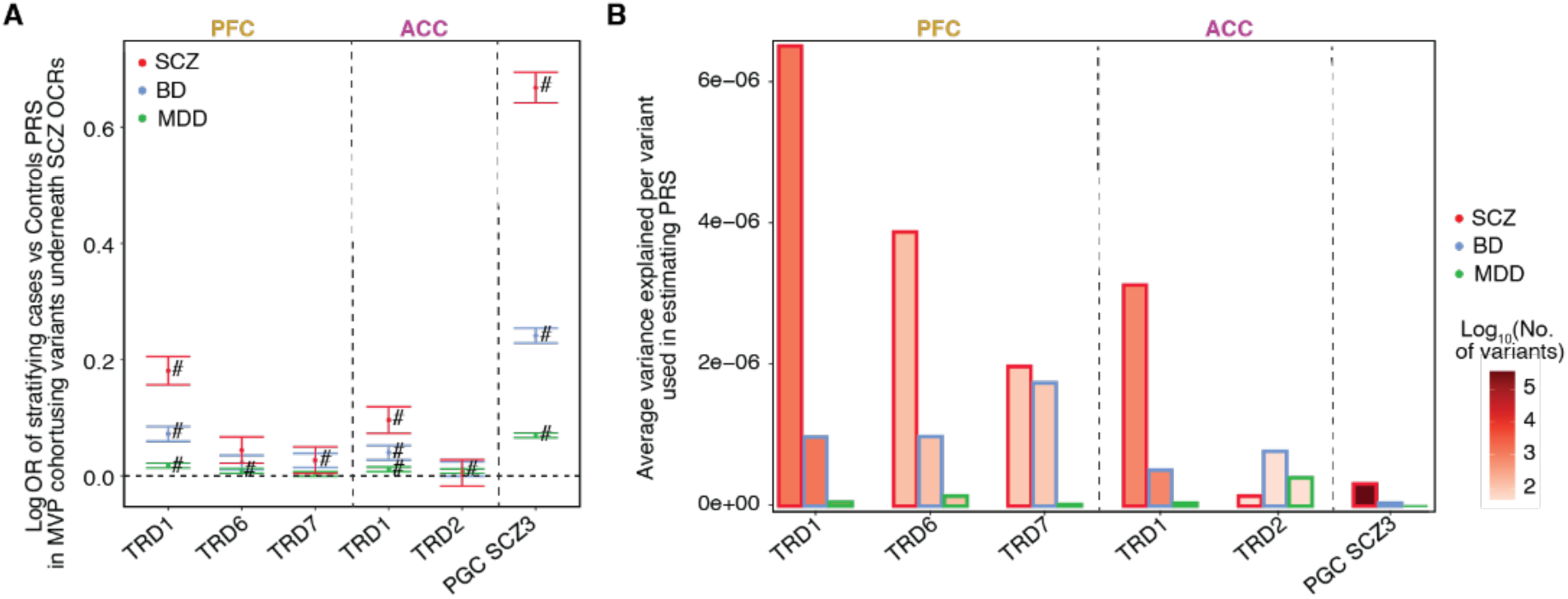
Trans Regulatory Domain one (TRD1) in PFC and ACC can stratify schizophrenia cases and controls in MVP cohort. For schizophrenia TRDs: TRD1, TRD6 and TRD7 from PFC region and TRD1, TRD2 from ACC region. **A**) Plot of logOR from logistic regression of PRS of 203,078 individuals from MVP cohort estimated using the SNPs in OCRs from these TRDs to predict their case-control status. Error bars show standard error in estimating OR from logistic regression. **B**) Barplot of the average variance explained per variant in estimating PRS of SCZ, BD and MDD individuals (bar contour color is red, blue and green respectively). Bar fill color corresponds to the number of variants considered in each group for PRS estimation (PFC TRDs: 1,388 for TRD1, 139 for TRD6 and 103 for TRD7; ACC TRDs: 815 for TRD1 and 46 for TRD2; PGC3 SCZ: 350,882).

## Discussion

We have created a large and comprehensive resource of 1,393 chromatin accessibility samples derived from neurons and non-neurons in the PFC and ACC of human brain specimens, including controls and cases with schizophrenia and BD. Differential chromatin accessible regions in neurons of schizophrenia brains were significantly associated with schizophrenia risk loci when compared to controls, which is consistent with a previous observation in the same cohort that perturbations in the enhancer-promoter-associated epigenetic marker, H3K27ac, are enriched in schizophrenia risk loci (*6*). Overall, these studies point to convergence among cell type-specific alterations in non-coding regions of the genome with the underlying genetic risk architecture of schizophrenia. We note the absence of results in BD due to less power in the sample size resulting in much lower numbers of associations with small effect sizes.

The concept of severity stages is well-established in diseases, such as AD (*27*), but has not been extensively explored in psychiatric disorders due to the absence of disease related molecular and pathological markers. A recent study in AD, applied the manifold learning method on bulk RNA-seq data from postmortem brains and identified severity stages that match disease stages based on pathological criteria (*20*). Using a similar approach, we derived a “disease pseudotime score”, stratifying samples into distinct clusters based on the extent of perturbation in the expression of disease-associated OCRs. The inferred order of samples was concordant with schizophrenia PRS across both cortical regions examined. This finding has the potential to open new avenues for the development of targeted drugs specific to different stages of the disease, similar to the concept of exploiting PRS for precision medicine in other disease fields (*28–30*).

In the next part of our study, we identified *cis*-regulatory domains (CRDs) of physically interacting chromatin regions, that constitute over 33% of genome-wide OCRs that are enriched for schizophrenia risk loci to CRDs only in neurons. To explore disease-relevant molecular mechanisms confined to higher order chromatin structures, we performed hierarchical clustering on disease associated CRDs based on their inter-individual correlation to identify trans regulatory domains (TRDs). Notably, one TRD (termed TRD1), which exhibited a high abundance of upregulated OCRs across both cortical regions, displayed a strong correlation with OCRs upregulated in fetal cortical samples compared to adult controls. By calculating the expression scores of schizophrenia OCRs from TRD1 in scATAC-seq data from fetal samples, we were able to pinpoint critical cell types in glutamatergic neurons at both early and late stages of development. The schizophrenia OCRs mapped to early stage glutamatergic neurons were related to DNA binding and translation, while schizophrenia OCRs mapped to late stage glutamatergic neurons showed processes related to synapse maturation and organization. These findings are concordant with the theory that non-coding regions, which host the majority of common schizophrenia risk variants, are linked to neurodevelopment.

Our findings lay the groundwork for characterizing neurodevelopmental cell types within diseased chromatin regions in adult postmortem brains that experienced disruptions during brain development. By gaining a deeper understanding of the roles played by neurodevelopmental cell types, we can establish connections between the molecular mechanisms of the disease and its manifestation during brain development, ultimately linking them to the symptoms observed in adulthood. Furthermore, there is compelling evidence supporting a neurodevelopmental basis for schizophrenia (*31–33*). Thus, this analysis provides a crucial link in interpreting the impact of risk mutations during the critical period of brain development, unravelling the susceptibility to schizophrenia. It should be noted that future improvements in the analysis could involve generating and analyzing scATAC-seq data from postmortem brains affected by schizophrenia and controls. This approach would allow for a more accurate mapping of disease-affected cell types to neurodevelopmental cell types, further enhancing our understanding of the condition.

Lastly, from this work, we provide unique resources of (1) dysregulated cell and region-specific chromatin regions in a cohort of schizophrenia and BD and (2) genome-wide CRDs that encompasses physically interacting OCRs and confines the schizophrenia risk loci from the rest of the OCRs. We also demonstrate (3) the stratification of schizophrenia samples based on the degree of perturbed epigenome. We (4) introduced a unique workflow based on hierarchical clustering of correlation of differential CRDs to map these neurodevelopmental OCRs to critical cell types using scATAC-seq data and (5) obtained a list of epigenomic marker OCRs that covers only 0.25-0.39% of risk SNPs and can stratify the cases and controls. The analyses and findings in this study provide the foundation for future investigations towards quantifying the contribution of specific cell types that are affected during brain development and later in life in brains of affected individuals. By understanding the extent of the contribution of specific cell types, we can gain valuable insights into the underlying mechanisms of the disease and potentially develop quantitative measures for assessing disease severity at a specific age, paving the way for personalized approaches to diagnosis and treatment in individual patients.

## Data Availability

All data produced in the present work are contained in the manuscript

## Acknowledgments

We thank the patients and families who donated material for these studies. Brain tissue for the study was obtained through the NIH Neurobiobank from the following brain bank collections: The Mount Sinai/JJ Peters VA Medical Center NIH Brain and Tissue Repository, Brain Tissue Donation Program at the University of Pittsburgh and the Human Brain Collection Core (HBCC) within the National Institute of Mental Health’s Intramural Research Program (NIMH-IRP). We thank the computational resources and staff expertise provided by the Scientific Computing at the Icahn School of Medicine at Mount Sinai. We thank members of the Roussos Lab for helpful discussion.

This research is based on data from the Million Veteran Program, Office of Research and Development, Veterans Health Administration, and was supported by award VA Merit I01BX004189. This publication does not represent the views of the Department of Veteran Affairs or the United States Government. We thank the participants of the Million Veteran Program, the scientists, clinicians and supportive staff involved in the construction of this biobank, and the scientific computing staff for the expertise that they provided.

## Author contributions

Conception and study design: P.R. Data generation: S.P.K, R.M, S.M.R and J.F.F. Data processing and analysis: KG., J.B., A.B, C.F., K.T., S.V., D.M., P.D., G.V. and G.E.H. Provision of brain tissue and resources: V.H. P.A. S.M. D.A.L. Writing of the paper: K.G., J.B. and J.F.F, P.R., with input from all authors.

## Data and materials availability

Raw (FASTQ files) and processed data (BigWig files, OCRs, and raw/normalized count matrices) has been deposited in synapse under synID https://www.synapse.org/#!Synapse:syn52264219/wiki/623312. Browsable UCSC genome browser tracks of our processed ATAC-seq data are available as a resource at: https://genome.ucsc.edu/s/girdhk01/CMC_ATAC. OCR coordinates, differential OCRS, CRDs, differential CRDs are available at the project repository at syn52264219.

## External validation sets used in the study are

ATAC-seq fetal specific peaks can be found at https://www.synapse.org/#!Synapse:syn52267265 and CTCF ChIP-seq on human neural cell (GEO GSE127577). TruSeq3-PE.fa file was downloaded from the adaptor folder under the trimmotic repository. https://github.com/timflutre/trimmomatic/blob/master/adapters/TruSeq3-PE.fa

The source data described in this manuscript are available via the PsychENCODE Knowledge Portal (https://psychencode.synapse.org/). The PsychENCODE Knowledge Portal is a platform for accessing data, analyses, and tools generated through grants funded by the National Institute of Mental Health (NIMH) PsychENCODE program. Data is available for general research use according to the following requirements for data access and data attribution: (https://psychencode.synapse.org/DataAccess).

## Additional information

Supplementary information is available for this paper at https://www.synapse.org/#!Synapse:syn52264219. Reprints and permissions information is available at www.nature.com/reprints. Correspondence and requests for materials should be addressed to P.R.

## Code availability

All publicly available software used is noted in the [Methods](https://www.nature.com/articles/s41593-022-01032-6#Sec10). We used decorate software to call CRDs: https://github.com/GabrielHoffman/decorate.

## Author Declarations

The Institutional Review Board (IRB) of University of Pittsburgh and Human Brain Collection Core and National Institute of Mental Health gave ethical approval for this work.

## Supplementary Information

This PDF file includes:

- Materials and methods
- Captions for Supplementary Figures S1-S21
- Supplementary Figs. 1-21
- Captions for Supplementary Tables S1-S13

Other Supplementary Materials/ Data Files for this manuscript include the following:

- Supplementary Tables 1-13 (.xlsx)

### Materials and Methods

#### Description of the post-mortem brain samples

Frozen brain tissue derived from ACC (anterior cingulate cortex/Broadmann area 10) and PFC (prefrontal cortex/Broadmann area 9 and 46) were obtained from three separate brain banks, described below. This cohort of schizophrenia (SCZ), bipolar or other affective/mood disorder (AFF) cases and control subjects was assembled after applying stringent inclusion/exclusion criteria. All involved subjects had to meet the appropriate diagnostic DSM-IV criteria, as determined in consensus conferences after review of medical records, direct clinical assessments, and interviews of family members or care providers. All tissue donors were from the Icahn School of Medicine at Mount Sinai (MSSM), University of Pittsburgh (PITT) brain banks and NIMH Human Brain Collection Core (HBCC). **Table S1A-D** tabulates the demographic information at sample level of the present study, including sex, age of death, and ethnicity, stratified by cell type, brain region, institution, and diagnosis. The link to the complete demographic and clinical information of the present study population is provided on synapse platform (syn52264219).

#### Generation of ATAC-seq libraries and sequencing

50mg of frozen brain tissue was homogenized in chilled lysis buffer (0.32M Sucrose, 5 mM CaCl_2_, 3 mM Magnesium acetate, 0.1 mM, EDTA, 10mM Tris-HCl, pH8, 1 mM DTT, 0.1% Triton X-100) and filtered through a 40µm cell strainer. Filtered lysate was underlaid with sucrose solution (1.8 M Sucrose, 3 mM Magnesium acetate, 1 mM DTT, 10 mM Tris-HCl, pH8) and subjected to ultracentrifugation at 24,000 rpm for 1 hour at 4°C. Pellets were re-suspended in 500µl DPBS supplemented with 0.1% BSA. anti-NeuN antibody (1:1000, Alexa488 conjugated, Millipore Cat #MAB377X) was added and samples incubated, in the dark, for 1hr at 4°C. Prior to FANS sorting, DAPI (Thermoscientific) was added to a final concentration of 1µg/ml. DAPI positive neuronal (NeuN+) and non-neuronal (NeuN-) nuclei were isolated using a FACSAria flow cytometer (BD Biosciences).

ATAC-seq libraries were generated using an established protocol (*1*). Briefly, 50,000 or 75,000 sorted nuclei were pelleted at 500 g  for 10 min at 4°C. Pellets were re-suspended in transposase reaction mix (22.5 μL Nuclease Free H_2_O, 25 μL 2x TD Buffer; Illumina Cat #FC-121-1030) and 2.5 μL Tn5 Transposase (Illumina Cat #FC-121-1030) on ice and the reactions incubated at 37°C for 30 min. Following incubation, samples were purified using the MinElute Reaction Cleanup kit (Qiagen Cat #28204, and libraries generated using the Nextera index kit; Illumina Cat #FC-121-1011) as previously described (*3*). Following amplification, libraries were resolved on 2% agarose gels and fragments ranging in size from 100-1000bp were excised and purified (Qiagen Minelute Gel Extraction Kit – Qiagen Cat#28604). Next, libraries were quantified by quantitative PCR (KAPA Biosystems Cat#KK4873) and library fragment sizes estimated using Tapestation D5000 ScreenTapes (Agilent technologies Cat# 5067-5588). ATAC-seq libraries were sequenced by Hi-Seq 2500 and Novaseq 6000 (Illumina) obtaining 50bp paired-end reads.

#### Processing of data

We have implemented our in-house pipeline for processing of ATAC-seq data previously used for other studies (*2*, *3*). The detailed steps are explained below.

##### Alignment of raw sequencing files

The raw reads were trimmed with Trimmomatic (*4*) and then mapped to human reference genome GRCh38 analysis set reference genome with the pseudoautosomal region masked on chromosome Y with the STAR aligner (v.2.7.0e) (*5*). To correct for allelic bias resulting from person-specific genome variation, we ran STAR with enabled WASP module (*6*) as we provided both ATAC-seq FASTQ file and WGS file or SNP array genotype of corresponding individual. This yielded for each sample a BAM file of mapped paired-end reads sorted by genomic coordinates. From these files, reads that mapped to multiple loci or to the mitochondrial genome were removed using samtools (*7*) and duplicated reads were removed with PICARD (v2.2.4; http://broadinstitute.github.io/picard). Quality control metrics were reported with phantompeakqualtools (*8*), ataqv (*9*), and Picard. All ATAC-seq QC metrics are summarized in **Table S1F** and details at sample level are provided on synapse platform (syn52264219).

##### Genotype calling

For ATAC-seq, genotypes were called using GATK (v3.5.0) (*10*). In brief, these steps were performed: (1) indel-realignment; (2) base score recalibration; and (3) joint genotype calling across all samples for variants with a phred-scaled confidence threshold >= 10. All clustered variants, variants in ENCODE blacklisted regions of the genome (*11*), and variants not in dbSNP v151 (*12*) were not considered. Read depth was not used for filtering. Finally, only variants with minor allele frequencies (MAF) ≥ 25% were considered. Pairwise genotype concordance was carried out amongst all ATAC-seq samples and whole genome sequencing (WGS) (or SNP-array samples for donors without WGS) using the kinship coefficient from KING v1.9 (*13*) and considering only the variants called in the ATAC-seq data. Cut-offs for the kinship values were selected based on the samples that were expected to be from the same brain versus those that were not. Using these cut-offs, we identified and resolved all sample swaps. We also reported potentially contaminated libraries that were matching multiple genotypes (or no genotype), mostly due to mistakenly used identical barcodes within the same pools for more than one sample. Those libraries were removed from the dataset.

##### Peak/OCR Calling

Peaks or OCRs calling was done as previously described (*2*, *3*) with a modification of FDR threshold to 0.01.

##### Metrics used for quality control

For each sample, the following quality control metrics were used: the total number of initial reads; the number of uniquely mapped reads; the fraction of reads that were uniquely mapped; further metrics from the STAR aligner; the GC content, duplication and insert metrics from Picard; the rate of reads mapping to the mitochondrial genome; the PCR bottleneck coefficient (PBC), which is an approximate measure of library complexity estimated as uniquely mapped non-redundant reads divided by the number uniquely mapped reads; TSS enrichment in housekeeping genes (calculated per ENCODE ATAC-seq data standards); the normalized strand cross-correlation coefficient (NSC) and the relative strand cross-correlation coefficient (RSC), which are metrics that use cross-correlation of stranded read density profiles to assess sample quality independently of peak calling; and finally the fraction of reads in peaks (FRiP), which is the fraction of reads that fall in detected peaks, the fraction of reads in only blacklisted peaks (*11*), and the ratio between these two metrics (to calculate these metrics, the consensus set of peaks was used). The main quality metrics are shown in **Table S1F** and details at sample level are provided on the synapse platform (syn52264219).

##### Annotation of OCRs to genomic elements

Neuronal and non-neuronal OCRs were annotated to TSS, exon, 5′ UTR, 3′ UTR, intronic or intergenic regions using ChIPSeeker (version 1.18.0). All OCRs within ±3 kb distance from TSS of a gene were annotate as promoters. **Fig. S5A** shows the distribution of neuronal and non non-neuronal OCRs annotated to (1) promoters, (2) introns, (3) distal intergenic and (4) exon and UTRs using the “TxDb.Hsapiens.UCSC.hg38.knownGene” transcript database in-built in the ChIPSeeker package.

#### Annotating OCRs to genes using ABC model

We used Activity-by-contact model (ABC, v.0.2) (*14*) to construct a comprehensive regulatory map of enhancer-promoter (E-P) interactions in neuronal and non-neuronal cell types of the two investigated brain regions (DLPFC and ACC). This model requires: (1) contact frequency between putative enhancers and promoters of regulated genes; and (2) enhancer activity data. Contact frequency matrices were generated from neuronal and non-neuronal Hi-C datasets composed of eight post-mortem human brains (*2*). Here different neocortical regions (dorsolateral prefrontal cortex, orbital frontal cortex, and anterior prefrontal cortex) were profiled across multiple donors aged 34-103 years. Enhancer activity data was represented by the cell type and brain region specific ATAC-seq signal (current study) and the H3K27ac ChIP-seq signal. ChIP-seq data was generated in a subset of ten controls (STG and EC; age of donors ranged between 61-103 years) (*15*). In accordance with the authors’ directions, we filtered out predictions for genes on chromosome Y and lowly expressed genes (genes that did not meet inclusion criteria in our RNA-seq dataset). We used the default threshold of ABC score (a minimum score of 0.02), the default screening window (5MB around the TSS of each gene), and we skip ABC calculation for ubiquitously expressed genes (using default ABC list of 806 genes).

#### Analysis of differentially accessible OCRs (schizophrenia OCRs)

To assess which OCRs showed differential accessibility in SCZ-and BD-related phenotypes, we analyzed the accessibility statistically. For this, chromatin accessibility was estimated by the number of ATAC-seq reads overlapping a given OCR. The more overlaps seen with an OCR the more accessible the more accessible the OCR was considered. The statistical analysis encompassed the following steps.

##### Read count and OCR filtering

The starting point here were the sample by OCR matrices (separately for neuronal and non-neuronal samples) of read counts generated as described in the preceding section. From these matrices, we excluded OCRs that were lowly accessible by only keeping OCRs that had at least 1 count per million reads in at least 10% of the samples. This removed 742 and 4 neuronal and non-neuronal OCRs, respectively, and resulted in a final read count matrices of 690 neuronal samples by 391,420 OCRs and 703 non-neuronal samples by 260,431 OCRs. Next, the read counts were normalized using the trimmed mean of M-values (TMM) method (*16*).

##### Exploration of covariates and model selection

To explore the effect of technical and biological covariates, we first did a principal component analysis (PCA) on the normalized read counts to identify high-variance components explaining more than 1% of the variance. We then accessed the correlation of covariates with the PCs and selected those that showed a significant correlation with one or more PCs at a lenient FDR cut-off of 0.2 as candidate covariates for the analysis of differential chromatin accessibility. This encompassed 64 covariates including FRiP, GC content metrics, mapping metrics, insert metrics, the fraction of reads mapping to the mitochondrial genome, PBC, RSC, and barcode. These covariates were subsequently assessed as detailed in the following.

Next, the starting point for modeling the chromatin accessibility was chosen as the variables “brain region by diagnosis status” (2x2=4 levels) and “sex” (2 levels) for a base model. The variable “sex” was included as it is known to have a strong effect on a few OCRs primarily located on the gonosomes. To assess which covariates should be included in order to have a good average model of OCR accessibility we employed the Bayesian information criterion (BIC). In particular, it was for each additional covariate tested how many OCRs showed an improved BIC score minus how many showed a worse BIC score when the covariate was included in the linear regression model compared to when it wasn’t. Here, a covariate was required to improve mean BIC per OCR by at least 10 in at least 2% of OCRs in order for it to be included in the final model. If there are some covariates fulfilling this criteria, the best performing of them is added into the base model and the remaining covariates are tested again. For neuronal dataset, the following numerical covariates were selected: “GC 80-100%” (i.e., normalized coverage over each quintile of GC content ranging from 80 – 100%), “Fraction of reads in peaks (FRiP)”, “Fraction of unmapped reads”, “GC 20-39%”, “AT dropout”, “GC 40-59%”, and “Age at the time of death”. For non-neuronal dataset, the following numerical covariates were selected: “GC 20-39%”, “GC 80-100%”, “GC 0-19%”, “Age at the time of death”, “Fraction of reads in blacklisted peaks (FRiBP)”.

Subsequently 14 categorical covariates were considered for inclusion due to the higher number of degrees of freedom of each covariate. None of these fulfilled the BIC criteria for inclusion. Finally it was considered if the selected numeric covariates affected chromatin accessibility as a quadric term by testing the squared FRiP for inclusion. It did not meet the BIC inclusion criteria and was therefore not added to the model. Resultantly the final neuronal and non-neuronal models jointly encompass 14 and 12 degrees of freedom, respectively (**Fig. S7**).

##### Statistical analysis of differences in chromatin accessibility

The normalized read count matrices from voomWithDreamWeights (variancePartition package (*17*, *18*)) were then modeled for each brain region (i.e.,PFC, ACC), and diagnosis (i.e., SCZ, BD, and Controls) by fitting weighted least-squares linear regression models estimating the effect of the right hand side variables on the expression/accessibility of each feature:

- neuronal OCR expression ∼ brain region:diagnosis + sex + GC_80-100% + FRiP + Fraction_unmapped_reads + GC_20-39% + AT_dropout + GC_40-59% + Age_of_death
- non-neuronal OCR expression ∼ brain region:diagnosis + sex + GC_20_39% + GC_80-100% + GC_0-19% + Age_of_death + FRiBP

Since our dataset contains three samples per individual, we ran differential analysis by *dream* method (variancePartition package (*17*, *18*)). *Dream* method properly model correlation structure and, thus, keep false discovery rate lower than the other commonly used methods for this purpose.

#### Analysis of differentially expressed genes

The analysis of differential gene expression follows the same protocol as previously described analysis of chromatin accessibility. Therefore, a description will focus on explanation of changes of parameters in the pipeline.

##### Read count and OCR filtering

The initial read count matrix of the CMC RNA-seq dataset (*19*) consisted of 58,930 genes quantified for 1,818 samples (only samples from DLPFC and ACC brain regions with SCZ, BD, and Control diagnosis status were selected). Then, we calculated the correlation of each sample to all other samples and removed the samples with markedly different correlation, i.e., the difference of mean correlation of the given sample with the rest of the dataset and mean correlation of all pairs of samples within the dataset was more than four times higher than the standard deviation calculated upon all pair’s correlation. We only kept protein-coding genes with gene expression of at least 1 count per million reads in at least 30% of the samples. These filters led to the exclusion of 66 samples and 44,024 genes so the resultant read count matrix consisted of 1,818 samples by 14,906 genes. We performed quantile normalization of the data followed by normalization using the trimmed mean of M-values (TMM) method (*16*).

##### Exploration of covariates and model selection

The following covariates were selected by BIC method to be added to the base covariates, i.e. “region by diagnosis status” (2x3=6 levels), “sex” (2 levels), and “institution” (4 levels), “Expression profiling efficiency” (numeric, squared), “Intragenic rate” (numeric, squared), “GC 80-100%” (numeric), “AT dropout” (numeric), “age at the time of death” (numeric), “GC 20-39%” (numeric), “intronic rate” (numeric), “GC 60-79%” (numeric), intergenic rate (numeric, squared), “duplication rate” (numeric). We required that at least 5% of the peaks showed a change of 2 in the BIC score, corresponding to “positive” evidence against the null hypothesis (*20*). The relatively high number of covariates can be explained by the study design which employed homogenate tissues, not FANS-sorted nuclei. Therefore, the model needed to cope with naturally different neuronal / non-neuronal composition in different brain regions. The final model jointly encompassed 24 degrees of freedom.

#### Identification of SCZ stages using manifold learning

To identify SCZ stages, neuronal and non-neuronal schizophrenia OCRs were first filtered based on FDR corrected p-values with thresholds of FDR < .0001 in PFC (n=2,411 neurons upregulated schizophrenia OCRs), FDR < .001 in ACC (n=1,495 neurons upregulated schizophrenia OCRs), FDR < .0001 in PFC (n= non-neurons upregulated schizophrenia OCRs), and FDR < .001 in ACC (n= non-neurons upregulated schizophrenia OCRs). Filtered expression matrices of cell-region specific OCRs were then projected onto the first 50 principle components and a nearest neighbour graph was constructed using the UMAP neighbour kernel (*21*). Diffusion maps (*22*) were calculated with the resulting neighbour graph using the kernel normalisation introduced in (*23*) (see **Fig. S11A-B**). We then constructed another UMAP neighbour graph using the first 15 diffusion coordinates and performed Leiden clustering (*24*) (resolution parameter = 1) to identify SCZ stages in diffusion map space. A total of 6(5) leiden clusters were obtained from PFC(ACC) regions in neuronal samples (see **Fig. 2C** and **S11C**). A diffusion pseudotime calculation was then performed according to (*23*) with a root node in the cluster with the highest density of control samples. All analysis was performed in scanpy using default parameters (see **Fig. S11D-E**).We limited the pseudotime estimation to neuron PFC and ACC samples only because of noisy diffusion maps for non-neurons PFC and ACC OCRs expression matrices.

The diffusion maps of the non-neuronal OCRs were notably noisier than those of the neuronal OCRs, leading to worse clustering and SCZ staging. One possible reason for this is the difference in spectral gaps, i.e. the difference between the eigenvalues associated with the first and second principal components of the data. When the data is Gaussian distributed, large spectral gaps lead to clean diffusion maps which follow the shapes shown in (see **Fig. S11A-B**), while smaller gaps lead to noisier ones. One biological reason for this is that the non-neuronal samples contain a mixture of cell types (astrocytes, oligodendrocytes, and microglia) as opposed to the neurons which only contain different subtypes. The first and second principal components of the mixture distribution can, in general, be orthogonal to the first and second principal components of each individual cell distribution and in such a case the spectral gap of the mixture distribution can be decreased and, in some cases, disappear entirely. If one cell type dominates the mixture distribution with a large number of samples and large variance, its principal components will dominate the mixture distribution while the other cell types will shrink the overall spectral gap of the mixture relative to the dominant cell type. This leads to decent diffusion maps for the first couple of diffusion components, followed by noisy diffusion components that deviate from the expected structure.

#### Polygenic Risk Scores

##### Million Veterans Program cohort

The Million Veteran Program (MVP) is a mega-biobank that has been previously described (*25–27*). On top of quality control performed by the MVP Core Team, we perform additional quality control (sex check, relatedness, minor allele frequency, hardy weinberg equilibrium, missingness, etc..) recommended for similar studies (*28*) using electronic medical data v20.1. For this study, we only utilise the European (EUR) population as defined by HARE(*29*) using the 1000 Genomes Project reference (*30*).

##### PRS estimation

Genotyping specifications for the CMC SNP array data used in the PRS analyses have been described previously(*19*, *31*). For the dataset used in these analyses specifically, marker positions were lifted-over to GRCh38. Pre-imputation processing then included running the quality control script HRC-1000G-check-bim.pl from the McCarthy Lab Group (https://www.well.ox.ac.uk/~wrayner/tools/), using the Trans-Omics for Precision Medicine (TOPMed) reference(*32*). Genotypes were then phased and imputed on the TOPMed Imputation Server (https://imputation.biodatacatalyst.nhlbi.nih.gov). Variants with an imputation R^2^ > 0.8 were retained. Samples with a mismatch between one’s self-reported and genetically inferred sex, suspected sex chromosome aneuploidies, high relatedness as defined by the KING kinship coefficient (*13*)(KING > 0.177), and outlier heterozygosity (+/- 3SD from mean) were removed. Additionally, samples with a sample-level missingness > 0.05 were removed, calculated within a subset of high-quality variants (variant-level missingness ≤ 0.02).

CMC samples of EUR ancestry, as defined by assignment to the EUR superpopulation described by the 1000 Genomes Project(*30*, *33*), were isolated using a 3SD ellipsoid method. Genotypes were first merged with GRCh38 v2a 1000 Genomes Project data (https://wellcomeopenresearch.org/articles/4-50) (*33*) using BCFtools version 1.9 (*34*). PLINK 2.0 (*35*) was then used to calculate the merged genotypes’ principal components (PCs), following filtering (minor allele frequency (MAF) ≥ 0.01, Hardy-Weinberg equilibrium (HWE) p-value ≥ 1 × 10^−10^, variant-level missingness ≤ 0.01, regions with high linkage disequilibrium (LD) removed) and LD pruning (window size = 1000 kb, step size = 10, r2 = 0.2) steps. An ellipsoid with a radius of 3SD, corresponding to the 1000 Genomes Project EUR superpopulation, was constructed using the first three genotype PCs. Only samples that fell within this ellipsoid (n = 923) were retained for subsequent variant-level filtering. Autosomal variants with an HWE p-value ≥ 1 × 10^−6^, MAF ≥ 0.01, and missingness ≤ 0.02 were thus retained, leaving 8,914,391 variants remaining.

SCZ PRS were calculated on the MVP and CMC cohort samples using summary statistics from the Psychiatric Genomics Consortium (PGC3) SCZ GWAS as training (*36*). PRS constructed with SNPs underneath schizophrenia OCRs within specific TRDs used as training data PGC3 SCZ GWAS summary statistics that were subsetted to include only SNPs falling within TRD1, TRD6, or TRD7 (for PFC-specific scores) and TRD1 or TRD2 (for ACC-specific scores), respectively. The PRS-CS-auto method (*37*) was used to apply continuous shrinkage priors to the effect sizes from these summary statistics.

A EUR LD reference panel provided by the developers of PRS-CS was utilized (https://github.com/getian107/PRScs), which draws from UK Biobank data (*38*). The following PRS-CS default settings were used: parameter a in the γ-γ prior = 1, parameter b in the γ-γ prior = 0.5, MCMC iterations = 1000, number of burn-in iterations = 500, and thinning of the Markov chain factor = 5. The global shrinkage parameter phi was set using a fully Bayesian determination method. Individual-level SCZ PRS and TRD-specific PRS were calculated using PLINK 2.0 (*35*). SCZ PRS and TRD-specific PRS were calculated in the MVP cohort similarly to how it was calculated above, except utilising a EUR LD reference panel drawing from 1000 Genomes Project data (*33*).

The associations between scaled (mean = 0, SD = 1) PRS and diagnostic status (SCZ vs. controls, BD vs. controls, MD vs. controls) were assessed using logistic regression. For current study samples, covariates included in the analyses were age at the time of death, age squared, the first 20 genotype PCs (computed within EUR ancestry samples only), sex, and institution (Mount Sinai, University of Pennsylvania, University of Pittsburgh, or NIMH Human Brain Collection Core). For the MVP cohort, we utilised the latest MVP Phenotypes (v21.1) and used covariates for age at recruitment, age squared, the first 20 genotypes PCs (computed within the whole cohort) and sex.

PRS regression coefficients were reported as the natural logarithm of odds (logOR) along with 95% confidence intervals, and all reported p-values are unadjusted. A variance partitioning tool (https://github.com/GabrielHoffman/misc_vp/blob/master/calcVarPart.R) was used to determine the variance in SCZ(BD, MD) diagnosis status explained by the PRS variable for each regression model. These values were then divided by the number of variants used to compute each PRS, respectively, to obtain the reported “Average variance explained per variant used in estimating PRS” metrics.

#### Cis regulatory landscape analysis

##### CRD calling

Cell-region specific covariates corrected expression matrices from four datasets were utilized for CRD analysis. The datasets included: 1) neuron PFC (391,420 OCRs X 291 samples), 2) neuron ACC (391,420 OCRs X 272 samples), 3) non-neuron PFC (260,431 X 299 samples), and 4) non-neuron ACC (260,431 X 278 samples). Since disease-specific changes were not observed in BD vs. control samples at the OCR level, the CRD analysis focused only on SCZ and control samples. The CRD calling pipeline followed the methodology described in Girdhar et.al (*39*). Briefly, the first step involved estimating hidden confounds using probabilistic estimation of expression residuals (PEER)(*40*) residualization on each of the four above-mentioned matrices separately. This was done to eliminate global noise and retain the local correlation structure of OCRs in physical proximity to each other. The retained OCR correlation structure facilitated their isolation using decorate software(*41*) in the next step.

Genome-wide CRDs were called using decorate software (version 1.0.14) with mean cluster sizes of domains (10, 25, 50, 80, 100) on 11 PEER-corrected matrices, PEERs{1, 5, 10, 15, 20, 25, 30, 35, 40, 45, 50}, for each of the four datasets separately. To filter out CRDs with weaker mean correlation values, a two-step process was implemented: 1) The OCRs per chromosome were permuted to create 10 permuted matrices. CRDs were called on all 10 permuted matrices. 2) The distribution of the mean absolute correlation (MAC) metric was created for both permuted matrices and original matrices separately, for the five cluster sizes (10, 25, 50, 80, 100). MAC is defined as the mean absolute correlation value of a CRD. We estimated the MAC cutoff by taking the MAC value at 95^th^ percentile from the distribution separately for each cluster size. **Figure S13** shows the plot of MAC vs PEERs for each cluster size. We choose PEER25 as optimal PEER for final CRDs all across the four datasets. All CRDs at the five mean cluster sizes were retained with a MAC value greater than MAC cutoff and any overlapping CRDs were merged into one after this step.

In total, 6,706(6,625) neuronal CRDs and 4,612(4,710) non-neuronal CRDs were obtained from PFC(ACC) regions (**Fig. 3B)** which encompassed, on average, of 21.7(22.1) and 18.9(18.7) OCRs per CRD that were present in neuronal and non-neuronal CRDs in PFC(ACC) respectively, with a median number of 6 OCRs in all four datasets (**Fig S15A** and **Table S4**). We estimated cell and region specificity of CRDs by measuring jaccard index across all pairs as shown in **Fig S15B**. To measure the odds of observing CRDs to be inside a subTADs, we ran Fisher test (*42*) by using 2 x 2 contingency table of OCRs within CRDs that are inside/outside subTADs in comparison to all OCRs that are inside/outside subTADs as shown in **Fig S15C**. To test whether the OCRs within the CRD contain three-dimensional genome interactions, we utilised CTCF ChIP-seq genomic regions from ENCODE human neural cells (*43*) (syn52264219) and quantified the density of CTCF sites in 200 bins (each bin size equals to 1 kb) around CRD boundaries (**Fig. S15D-G)** and subTAD boundaries from HiC datasets for the reference (**Fig. S15H**).

Alternatively, we measured the odds of enhancer-promoter OCRs from the ABC model within a CRD vs all OCRs in a CRD. To do this, we ran Fisher test (*42*) by using 2 x 2 contingency table of cell-region specific OCRs from ABC model (*14*) that are inside/outside cell-region specific CRDs in comparison to all cell specific OCRs that are inside/outside CRDs as shown in **Fig S15I**.

#### Identification of schizophrenia CRDs

In our analysis, we employed a two-stage testing approach using the stageR package (*44*) to identify significant CRDs. The process involved two distinct stages: the screening stage (stage 1) and the confirmation stage (stage 2). In the screening stage, we summarized each CRD by computing its p-value using the Sidak method(*45*), which was applied to all p-values of OCRs within that particular CRD. In confirmation stage (stage 2), we focused only on CRDs with Sidak p-values less than 0.05 from stage 1. These selected CRDs underwent individual hypothesis testing to assess the dysregulation of OCRs within them. A CRD was considered a “schizophrenia CRD” if it obtained a p-value < 0.05 in stage 2, indicating significant differences in OCR expression between the SCZ and control samples. In order to summarize the directionality of CRDs, we calculated the mean of log_2_FC (SCZ vs Controls) of OCRs within each CRD, comparing SCZ samples to control samples. An upregulated CRD had a mean of log_2_FC (SCZ vs Controls) > 0, while a downregulated CRD had a mean of log_2_FC (SCZ vs Controls) <0 as depicted in **Fig S14C**. Differential CRDs summary table of p-values obtained from both stage 1 and stage 2 for all the identified CRDs across the four datasets is available at syn52264219.

#### Assessing the link of CRDs with schizophrenia OCRs and diseased genes

In order to assess whether the diseased OCRs are more likely to be within the CRD compared to outside the CRD, we employed a generalized linear model using logistic regression. The model used the t-statistics of OCRs from differential OCRs table at syn52264219 to predict the status of OCRs inside or outside the CRD as shown in eq(1). **Figure S16B** shows the odds of OCRs inside CRD from eq(1).

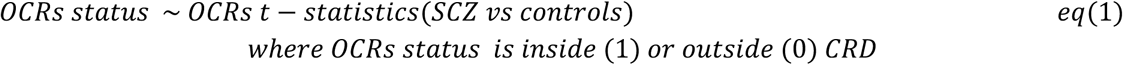

Subsequently, to examine whether the schizophrenia CRDs are indeed associated with schizophrenia OCRs, we utilized eq(2). This model aimed to predict whether the CRDs is schizophrenia CRD or not using the proportion of schizophrenia OCRs inside CRDs. We have added an offset term to control for the number of OCRs within the CRD. The prediction uses Poisson model distribution in *glm* R function. **Figure S16C** shows the odds of observing the clustered schizophrenia OCRs in SCZ CRD.

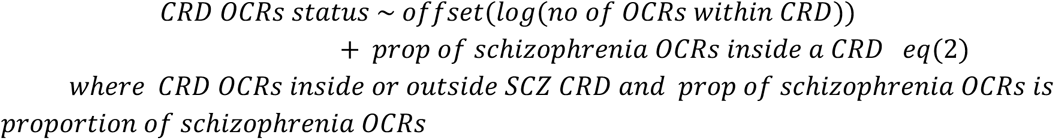

Lastly, to establish a connection between the disease-associated epigenome and the disease-associated transcriptome, we tested whether schizophrenia OCRs inside the schizophrenia CRDs could be predicted using the t-statistics of SCZ associated genes using eq(3). We use differential genes analysis from the CMC consortium (*19*). The OCRs were mapped to genes using ABC mapped genes table at syn52264219. **Figure S16E** shows the odds of observing the schizophrenia OCRs inside CRDs using the t-stats of differential genes.

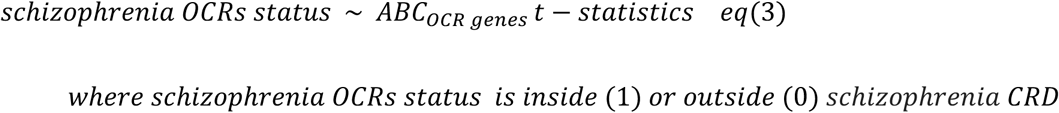

#### Clustering of schizophrenia CRDs into TRDs

In this section, we evaluate whether the interaction between schizophrenia CRDs across samples can stratify the CRDs into domains called trans-regulatory domains (TRDs) that can inform us about the specific molecular mechanism. We have limited the identification of TRDs to only schizophrenia CRDs in neuron PFC and neuron ACC because no significant association of CRDs with SCZ risk loci was observed in non-neurons ACC and PFC CRDs. The workflow from schizophrenia CRDs to TRDs is explained in schematic in **Fig. S17A**.

First, to obtain the interaction between schizophrenia CRDs, we took covariate corrected matrices of OCRs within neuron PFC and neuron ACC with dimensions of 1,953 x 360 and 1,691 x 330 respectively. Every CRD is summarized by calculating the average expression of OCRs using the eq(4) below:

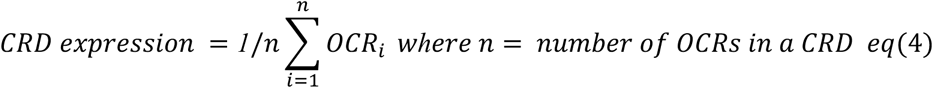

We took the Pearson correlation of CRD expression matrices that gives 1,953 x 1,953 and 1,691 x 1,691 dimensions of neuron PFC and ACC respectively. We used the first three principle components of correlation matrices for running clustering algorithm. We then fitted these matrices to hierarchical clustering with k=2:20 using diceR package in R using the default method of distance i.e. average linkage (*46*).

For every value of *k*, a random subsampling of 80% of the CRD correlation matrix is carried out 20 times. Therefore, not every sample is included in each clustering. The clustering for each iteration of the hierarchical clustering is completed using k-nearest neighbor (*47*) and majority voting. The optimal number of clusters was found by evaluating Baker-Hubert GAMMA index (*48*) while varying the cluster size (from *k*=2:20). GAMMA index is a measure of compactness (how similar are the objects within the same cluster), separation (how distinct are objects from different clusters), and robustness (how reproducible are the clusters in other datasets). Using GAMMA index, we found *k*=10 optimal clusters across both cortical regions for neuron CRDs as shown in **Fig. S17B**. The list of annotated TRDs to CRDs from the hierarchical clustering at *k*=10 clusters is available at syn52264219. For every CRD, we estimated its directionality using eq (5). We added these annotations of TRDs and their direction from eq (5) to the heatmap of correlation matrix of schizophrenia CRDs in **Fig 3D** and **Fig. S18A.** Also, we curated two lists, one with all OCRs and second one with only schizophrenia OCRs within each TRDs for all downstream analysis. **Fig. S17C** shows the number of all OCRs and schizophrenia OCRs within each TRDs whereas **Fig. S17D** is stratified by directionally of SCZ OCRs within TRDs. For **Fig. 3H** and **Fig. S18E,** we took all OCRs within TRD1 to measure the spearman correlation with fetal associated changes detected at FDR < .05 in Rahman et.al. (*49*). For the full differential OCRs table of Fetal cortical ATAC OCRs vs. Adult controls OCR analysis, see syn52264219.

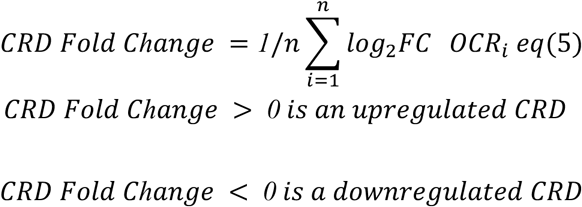

#### Fetal cortical ATACseq

##### Sample collection

Fetal brain samples were obtained from cortical plates from four distinct donors aged 18-24 pcw. These samples were collected from de-identified prenatal autopsy specimens with no neuropathological abnormalities at the Icahn School of Medicine at Mount Sinai. Library prep and post processing of atac libraries was done similar to described above.

#### Association test of fetal cell types to TRDs

We obtained scATAC-seq expression data from human fetal cortical samples, as outlined in the study by Trevino et.al (*50*). The dataset comprises four samples collected over an 8-week period during mid-gestation, specifically at 16pcw, 20pcw, 21pcw, and 24pcw, containing 6423, 4486, 12675 and 7720 cells. This dataset contains 31,304 nuclei annotated to a total of 22 cell-type clusters, including five types of interneurons (IN), nine different clusters of cortical excitatory neurons (GluN) and precursor cells, such as early radial glia, late radial glia and other non-neuronal cell types. In total, there were 657,930 fetal OCRs genomic regions identified in this dataset. Only the cell types with at least 100 cells in the cell type cluster were considered for this analysis.

To assess the expression score of schizophrenia OCRs within TRDs in fetal cortical cell types, we compiled a list of schizophrenia OCRs from both TRDs and outside TRDs that overlapped at least 99% with the fetal OCRs in both cortical regions. As a result, we obtained the majority of regions for TRD1 and outside TRDs, with neuron TRD1 comprising 3,189(2,490) regions from the PFC(ACC), while outside TRDs encompassed 2,055(1,383) regions from PFC(ACC). In contrast, the number of regions that overlapped with schizophrenia OCRs in TRDs was limited, with only 263(88) regions identified in neuron PFC(ACC) TRDs(2:10). To analyze the data further, we utilized the clustering annotations provided by Trevino et.al and computed the expression scores per cluster/cell types using Seurat’s AddModuleScore function. For better visualization, we did Z-score normalization on these module scores and generated plots, as shown in **Fig 4A** and **Fig S19A**.

To investigate significant cell types associated with expression scores from our input list, our approach was two fold. First we identified TRDs that exhibited a positive and significant association with a particular cell type, relative to other cell types. To achieve this, we applied a linear mixed model, considering the enrichment scores for each TRD at different developmental stages (pcw16, pcw20, pcw21, pcw24). The model was formulated as follows: (enrichment scores ∼ cell type + (1|sample ID)), where enrichment scores were obtained from the *AddModuleScore* R function(*51*). The cell type was coded as “cell type “ for cells in a given cluster and “non_celltype” for cells in other clusters, and sample ID represented the individual ID of cortical samples. By incorporating the sample ID as a random effect in the model, we accounted for the presence of multiple cells from each individual. We ran this model on all cell type scores to examine their associations. Finally, we created the list of cell types and associated TRDs that had significant and positive association after FDR correction for multiple testing burden from the number of tests.

In the second step, we ran another model to compare the enrichment scores of a given cell type from a significantly associated TRD with the enrichment scores of the same cell type from schizophrenia OCRs outside the TRD. The model was defined as lmer(enrichment scores ∼ TRD_OCRs_status + (1|Sample.ID) + (1|Cell.ID)), where enrichment scores were obtained as previously explained, and TRD_OCRs_status was coded as “TRD” for enrichment scores of cells from a significant TRD in a given cell type and “outside_TRD” for cells from outside TRD in the same cell type. The sample ID and cell ID represented the individual ID and cell IDs of cortical samples, respectively. We incorporated the cell ID and sample ID as a random effect to account for the repeated measurements from individuals and cells from the outside TRD enrichment scores. **Table S7A-B** and **Table S7C-D** show the output from step1 and step 2 on neuron PFC(ACC) TRDs. **Fig. 4B-C** and **Fig S19B-C** show the distribution of enrichment scores coloured by the magnitude of log_2_FC which is the enrichment score of schizophrenia OCRs in TRD1 in a given cell type vs enrichment score schizophrenia OCRs in TRD1 in other cell types from step 1. The star (*) annotation next to distribution shows whether or not a given cell type is significant after step 2. Our analysis showed significant association of schizophrenia OCRs in TRD1 with early glutamatergic cell types: Glu(1,3,5,8) and late glutamatergic cell types Glu(6,9) across both cortical regions. In inhibitory neuron class, IN(3,4,5) and IN(2,3,4,5) significant association with schizophrenia OCRs in TRD1 from PFC and ACC respectively. Next we gathered the cell specific markers from (*50*) for all significant cell types and intersect with ABC schizophrenia OCR mapped genes in TRD1 to obtain the cell-region and disease specific genes (see **Table S7G-H**). We used this list for running magma and pathway analysis as explained below to generate **Fig 4D-E** and **Fig. S19D-E.**

#### LD score enrichment analysis of CRDs

We ran LD score enrichment analysis to estimate the enrichment of brain-related and non-brain-related GWAS in all input OCR regions. For **Fig 2**, we did this analysis on all neuronal and non-neuronal OCRs using differential OCRs table (syn52264219), schizophrenia OCRs from neuron PFC, neuron ACC, non-neuron PFC and non-neuron ACC given (syn52264219). For **Fig 3**, we estimated SCZ heritability using schizophrenia OCRs within neuron PFC and neuron ACC CRDs, and outside neuron PFC and neuron ACC CRDs given in **Table S3, S6, S8.** In **Fig 4**, we did this analysis on OCRs within TRD1:10 from neuron PFC and ACC using **Table S7.** For all analyses, the broad MHC-region (chr6:25-35MB) was excluded due to its extensive and complex LD structure, but otherwise default parameters were used for the algorithm.

#### MAGMA association trait analysis

We used MAGMA version 1.08b (*52*) for all the analysis described below. For **Fig. 4E-F** and **Fig. S19E-F** we took the gene markers list obtained from the above section named as “Association test of scATAC from fetal cell types to TRDs’’**. Table S7G-H** tabulates cell-region and disease specific genes markers that were utilized to identify enrichment of GWAS brain and non-brain related traits in cell-region and disease specific genes. For each gene and trait, MAGMA calculates gene-level P-value based on the joint association of all SNPs from summary statistics that fall to the gene region while it accounts for linkage disequilibrium (LD) between SNPs. The gene regions were defined with the window size of 35kb upstream and 10kb downstream and LD was estimated from the European panel of 1000 Genome Project phase 3 (*33*). Then, MAGMA uses a linear regression framework to test whether the differentially expressed genes are more significantly associated with GWAS traits compared to the rest of the genome. In concordance with ATAC-seq GWAS analysis, we excluded genes that overlap the MHC-region(chr6:25-35MB).

#### Pathway analysis of OCRs

To functionally interpret the OCRs within TRD1 that overlapped with fetal specific OCRs (*49*) we used the Genomic Regions Enrichment of Annotations Tool (GREAT) approach to identify the biological function of nearby genes for regions using GREAT as shown in **Fig. 4F** and **Fig. S19F**. To interpret the biological function of cell-region and disease specific genes from section “Association test of scATAC from fetal cell types to TRDs’’, we applied GREAT pipeline to obtain **Fig 4D**, **Fig S19D** using the **Table S7G** and **Table S7H** respectively.

### Supplementary Figures

**Figure S1.**
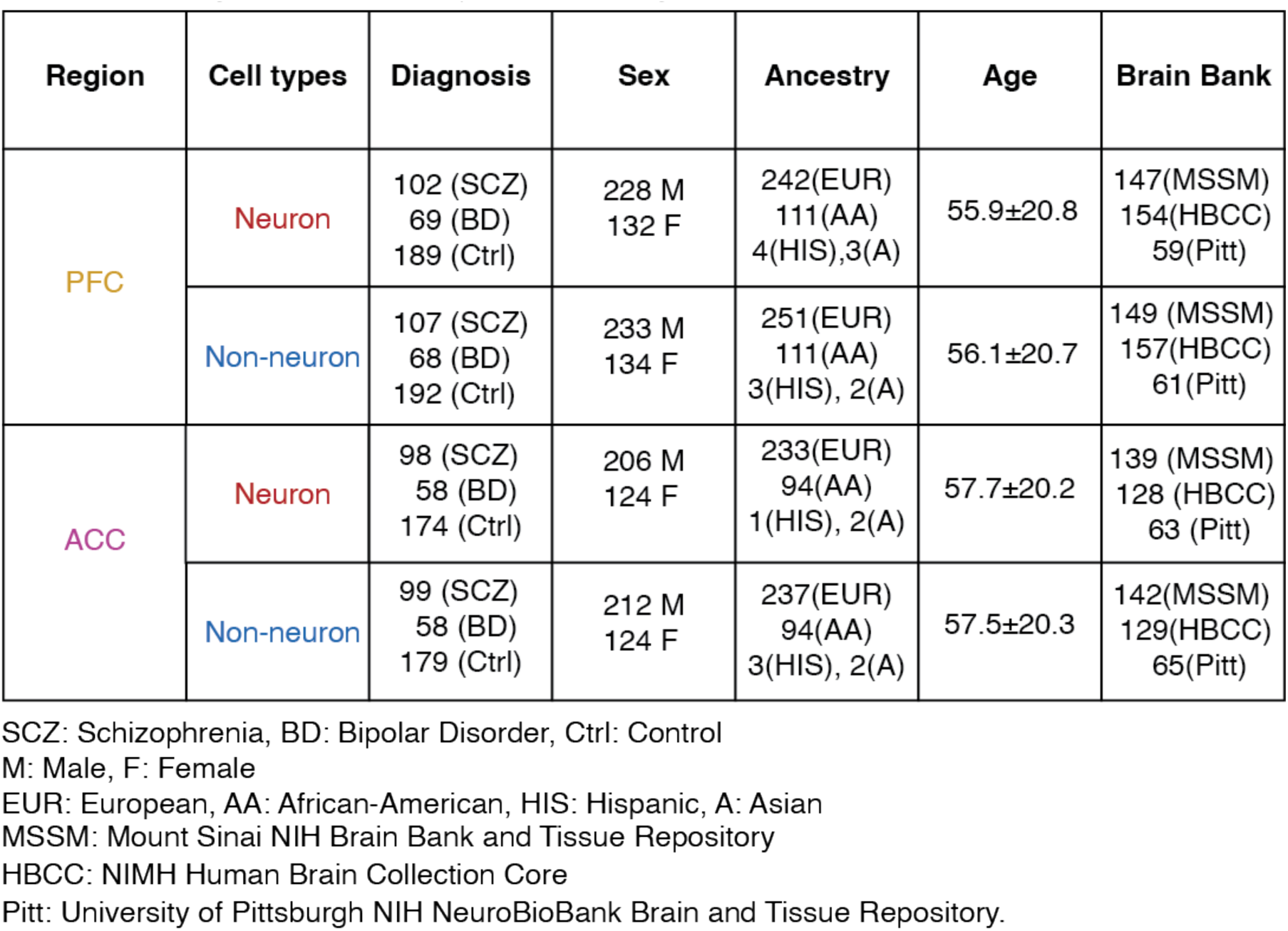
Summary metadata of samples in the study.

**Figure S2.**
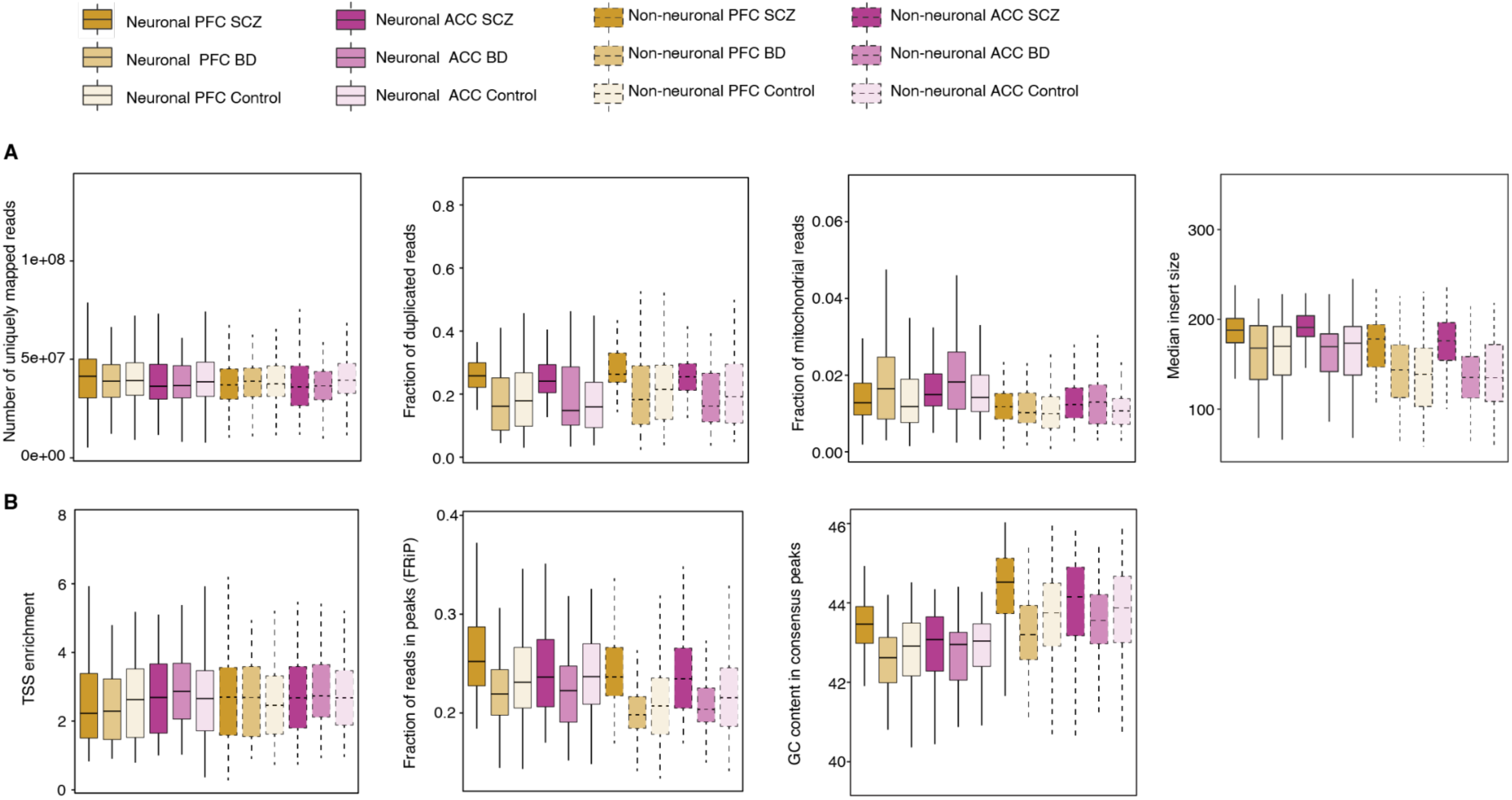
Quality control metrics in cell- and region-specific samples stratified by diagnosis. **A)** Distribution of uniquely mapped reads, fraction of duplicated reads, fraction of mitochondrial reads, and median insert size for each sample. **B)** Distribution of TSS enrichment of OCRs using all housekeeping genes from Refseq (*9*), fraction of reads in OCRs, and gc content in OCRs for each sample. In both A) and B), box plots are depicted with the center line representing the median, the box indicating the interquartile range (between the 25th and 75th percentile), and whiskers extending to the lowest and highest values within 1.5 times the interquartile range from the median.

**Figure S3.**
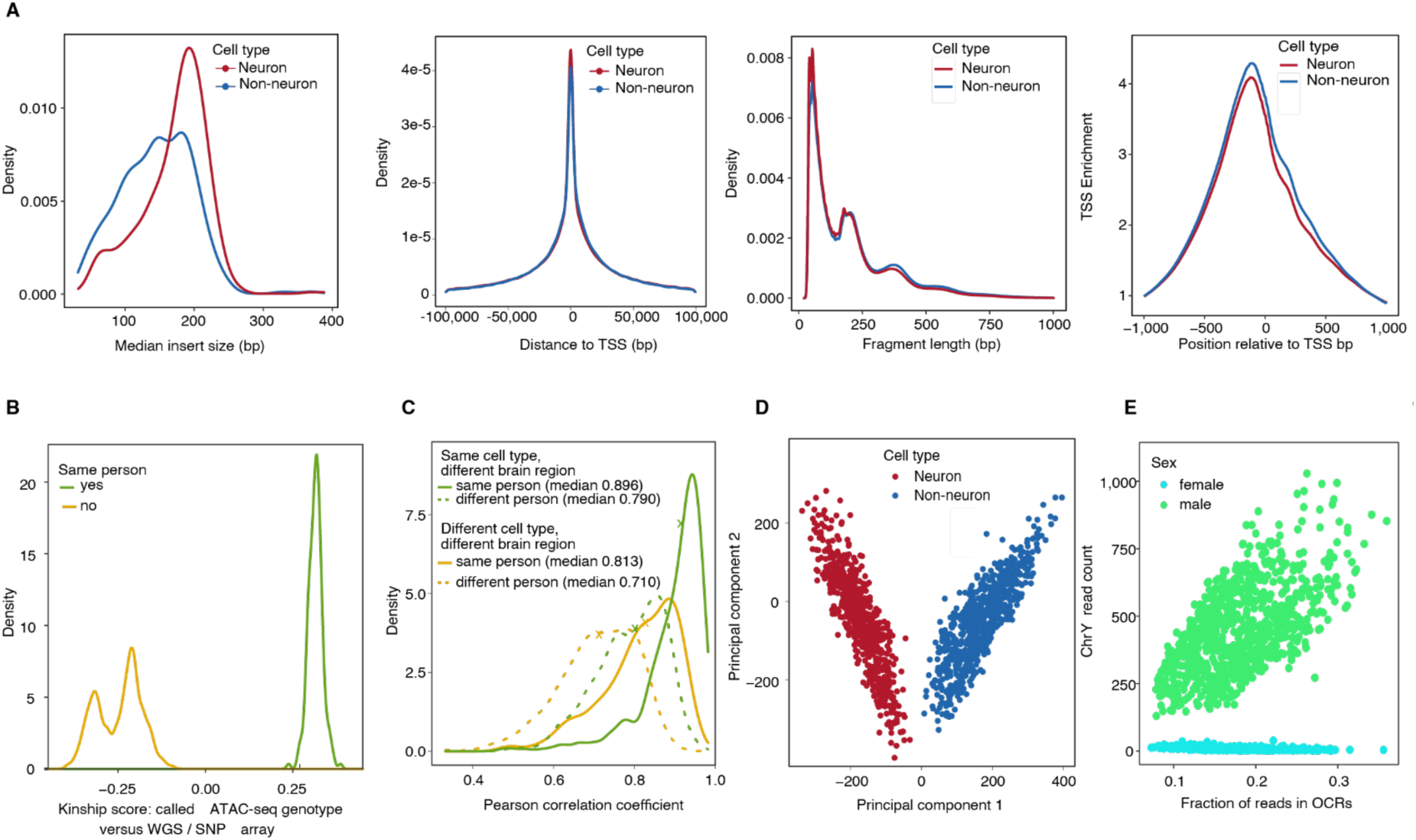
Quality control metrics of neuronal and non-neuronal consensus OCRs. **A)** Distribution of median read insert size, distance to TSS, fragment length, and TSS enrichment of cell-specific consensus OCRs. **B)** Genotype checks were performed using pairwise comparisons of genotypes called from ATAC-seq with genotypes from whole-genome sequencing, as well as with the ATAC-seq libraries themselves. **C)** Correlations of raw reads coverage (number of reads) over consecutive bins of 10 kb genomic regions between samples originating from the same versus different person (yellow line: samples originating from the same cell type but different brain region; green line: samples originating from different cell types but the same brain regions). Median correlations are highlighted by the “x” symbol.” **D)** Principal component analysis (PCA) was applied to evaluate chromatin accessibility levels in OCRs, with cell types represented in red for neuronal and blue for non-neuronal cells. **E**) Sex check based on measuring the number reads mapped on chromosome Y.

**Figure S4.**
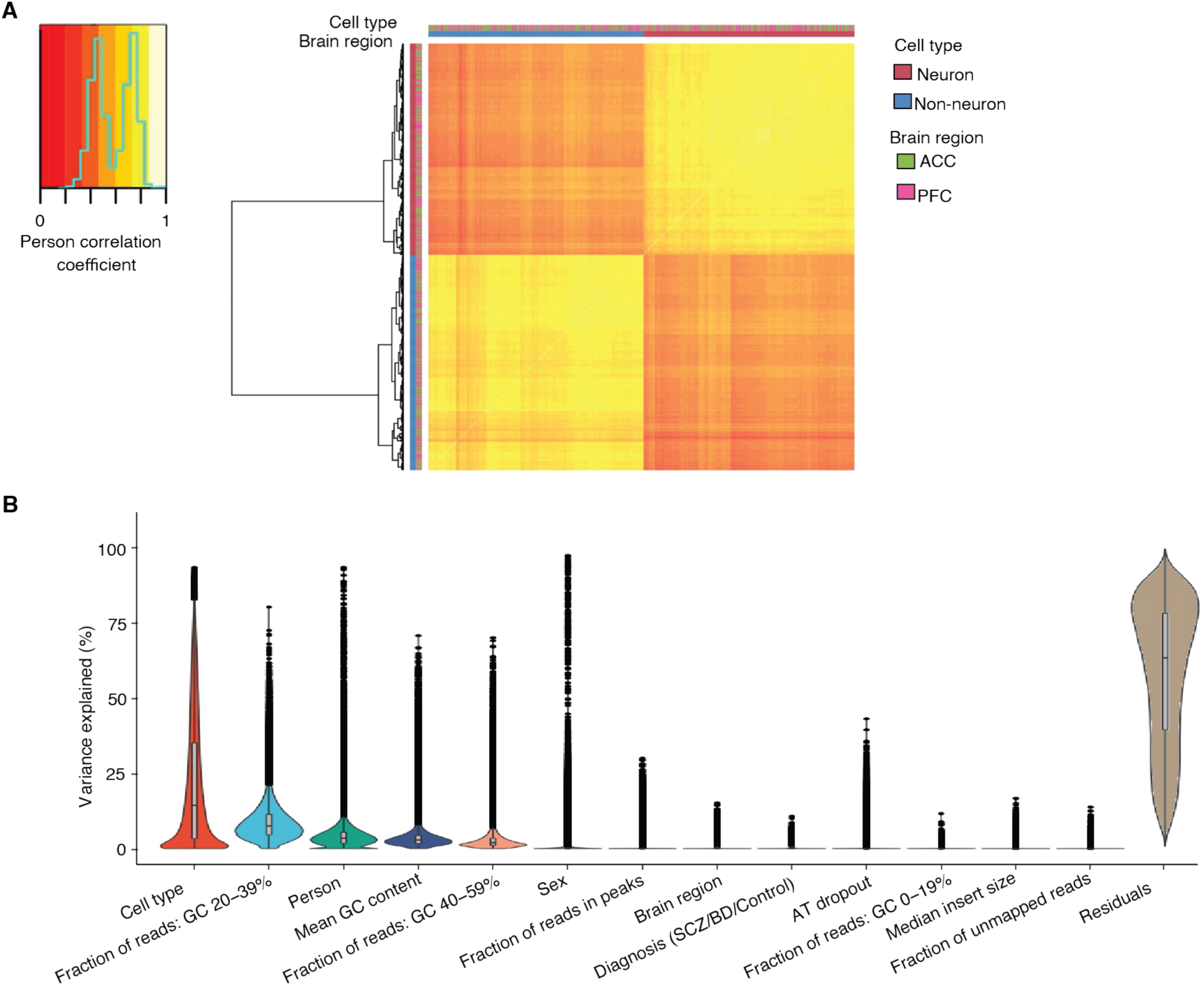
Variance explained by identified covariates in normalized counts of samples for all OCRs. **A)** Heatmap representing the Pearson correlation of expression levels for merged chromatin regions across all samples. The bar above the heatmap displays cell and region type labels corresponding to the samples. **B)** Distribution of variance explained by each OCR with respect to different covariates. The covariates on the x-axis were selected using a BIC model explained in the methods section. Box plots are depicted with the center line indicating the median, the box representing the interquartile range (between the 25th and 75th percentiles), and whiskers extending to the lowest and highest values within 1.5 times the interquartile range from the median. Dots are used to indicate potential outliers beyond this range.

**Figure S5.**
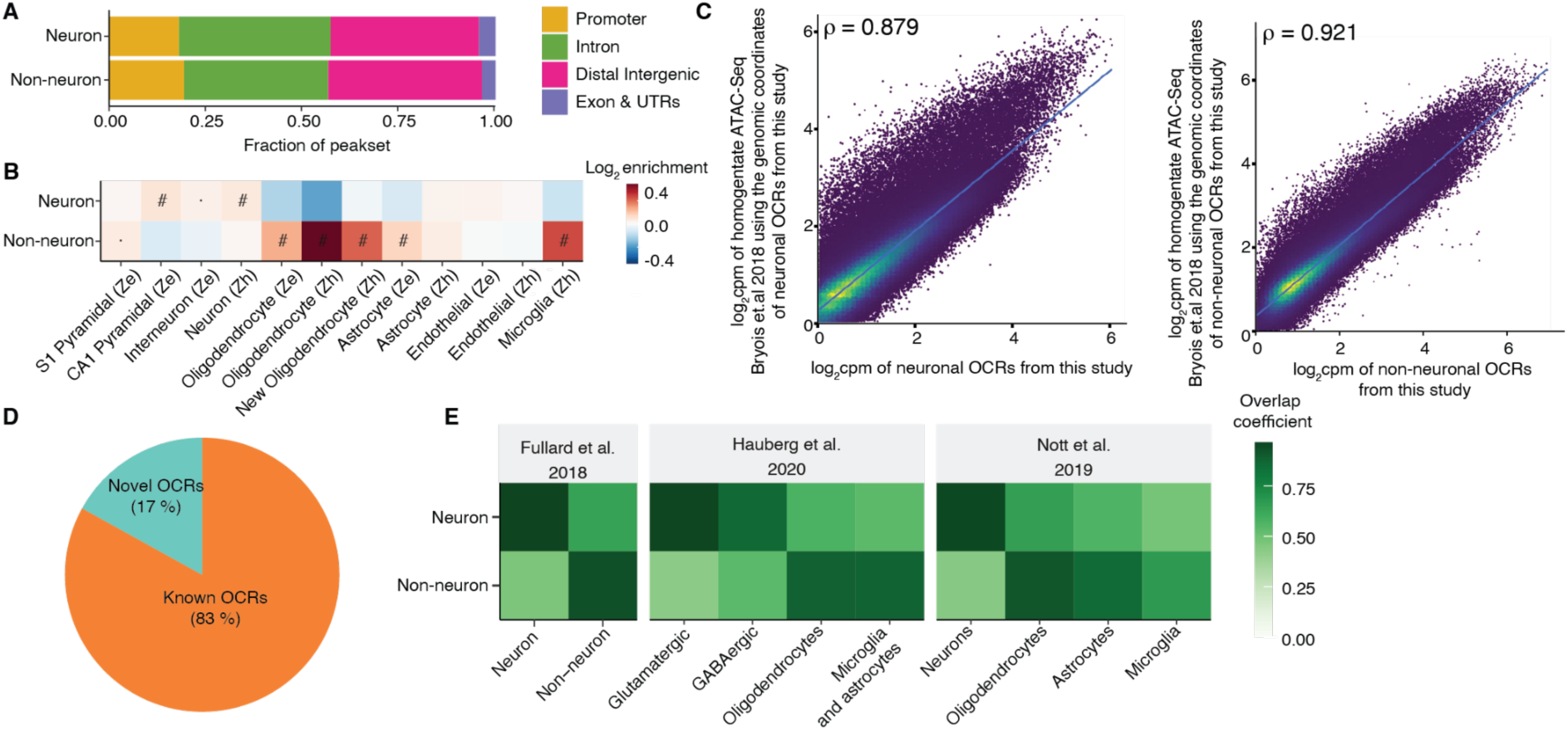
Annotation of OCRs to regulatory elements and similarity of OCRs with other datasets. **A)** Bar plot depicting the percentage of regulatory elements in each cell-specific OCR, obtained after annotation using ChIPseeker. **B)** Enrichment analysis showing the overlap of cell-specific OCRs with cell-specific marker genes using the resource from ‘Ze’ (Zeisel)(*53*) and ‘Zh’ (Zhang)(*54*). ‘#’: significant for enrichment after FDR correction of multiple testing across all tests in the plot (Benjamini–Hochberg test); ‘·’: nominally significant for enrichment. **C)** Spearman correlation analysis illustrates the expression count (log_2_cpm) correlation of neuronal OCRs from samples in this study with expression counts (log_2_cpm) of samples from PFC homogenate in Bryois et al.’s (*55*) study. **D)** Pie plot displaying the distribution of OCRs categorized as known (overlapping with other studies(*2*, *3*, *55–59*)) and novel (unique to this study). **E)** Overlap coefficient to show the overlap of OCRs identified in this study with cell-specific OCRs from other published datasets(*3*, *56*, *57*).

**Figure S6.**
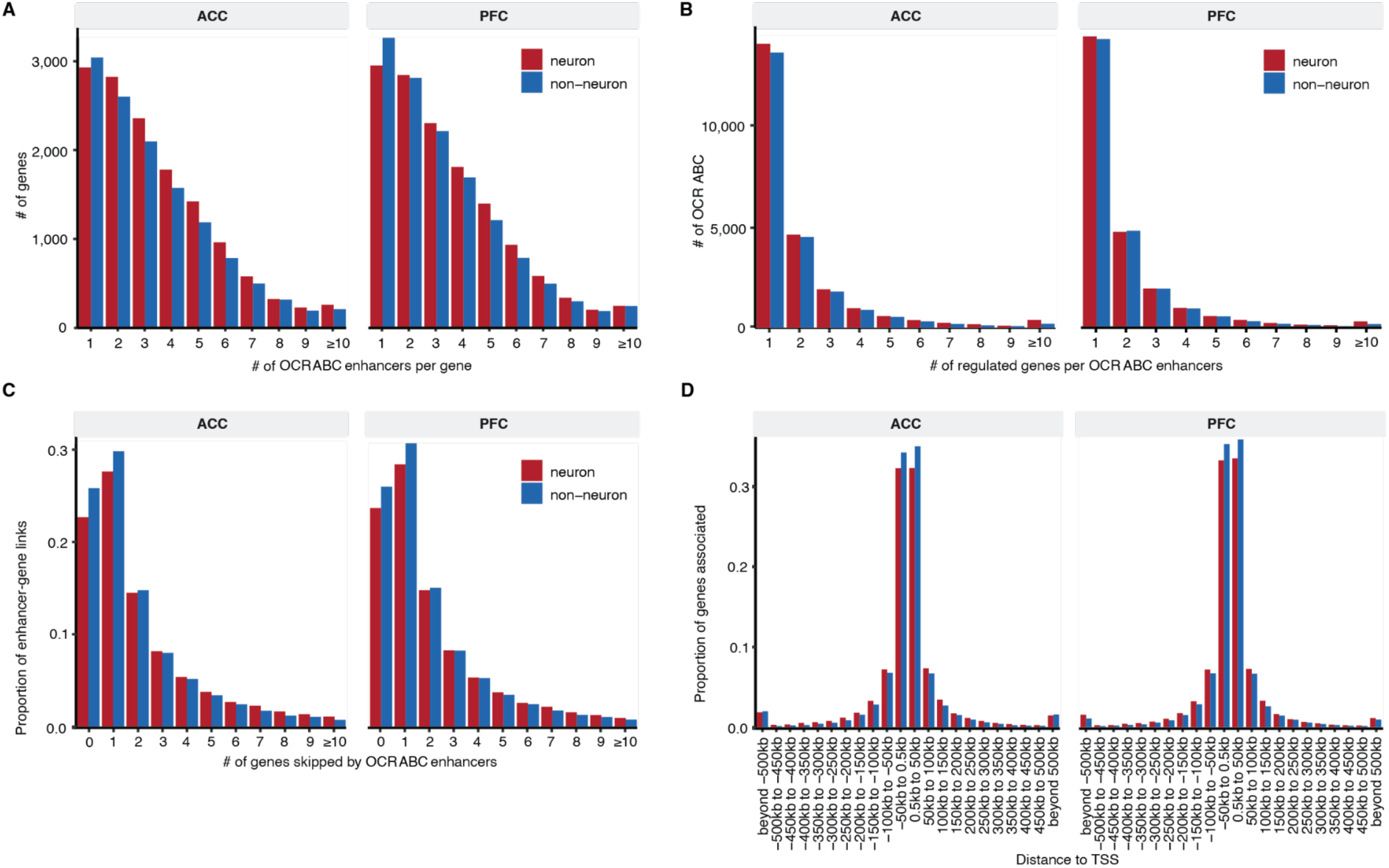
Annotation of OCRs to genes using the Activity-By-Contact (ABC) method. **A**) Histogram of the number of *OCR_ABC_* linked per gene. **B**) Histogram of the number of genes linked per *OCR_ABC_*. **C**) Histogram of the number of genes “skipped” by an *OCR_ABC_* to reach their linked genes. **D**) Histogram of the distance of *OCR_ABC_* to the TSS of regulated genes.

**Figure S7.**
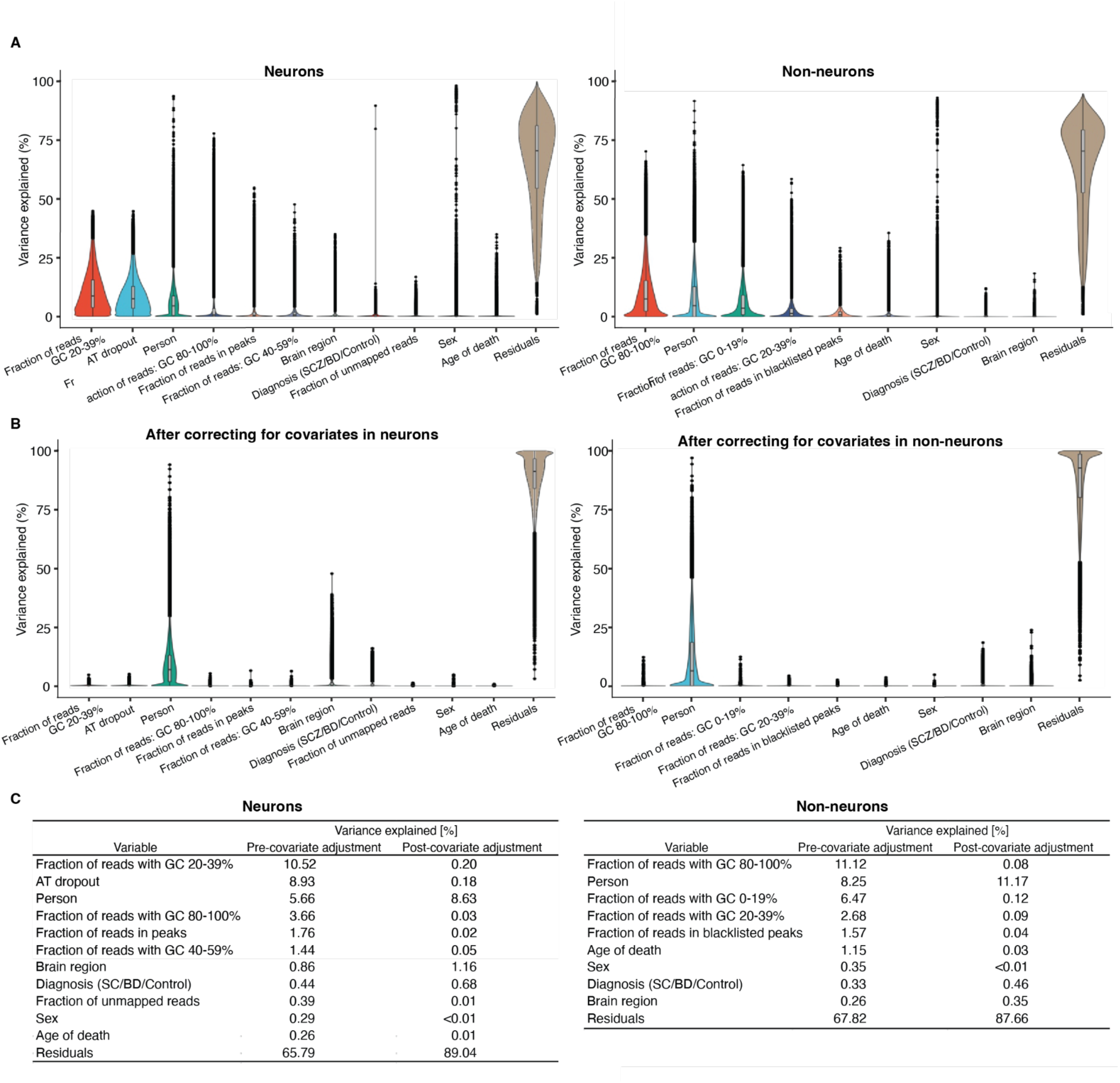
Covariate correction of the expression of cell specific OCRs. **A)** Violin plots to show the distribution of variation of OCRs for biological and technical covariates in neurons (left) and non-neurons (right) before and **B)** after correcting for covariates. All technical covariates (i.e. except Sex, Brain region, Person and Residuals) were identified using the BIC model explained in the methods section. **C)** Mean proportion of overall variance in OCRs attributed to the biological and technical covariates before and after correcting for technical (but not biological) covariates in neurons (left) and non-neurons.

**Figure S8.**
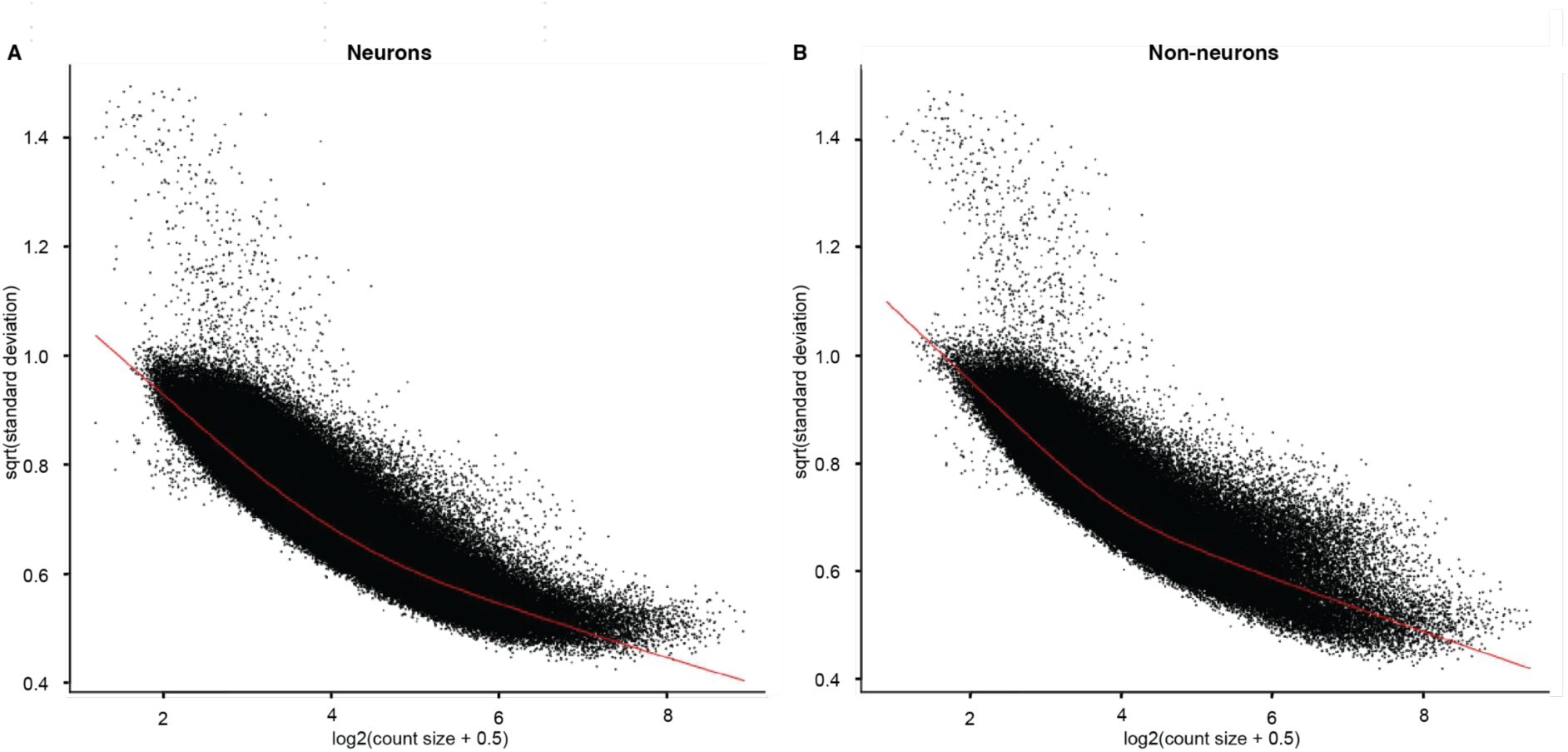
Mean variance distribution of normalized counts of neuronal and non-neuronal OCRs. **A)** and **B)** show the plot of mean vs variance in expression of neuronal and non-neuronal OCRs shown as black dots with LOWESS trend as a red line that indicate low -moderate biological variation.

**Figure S9.**
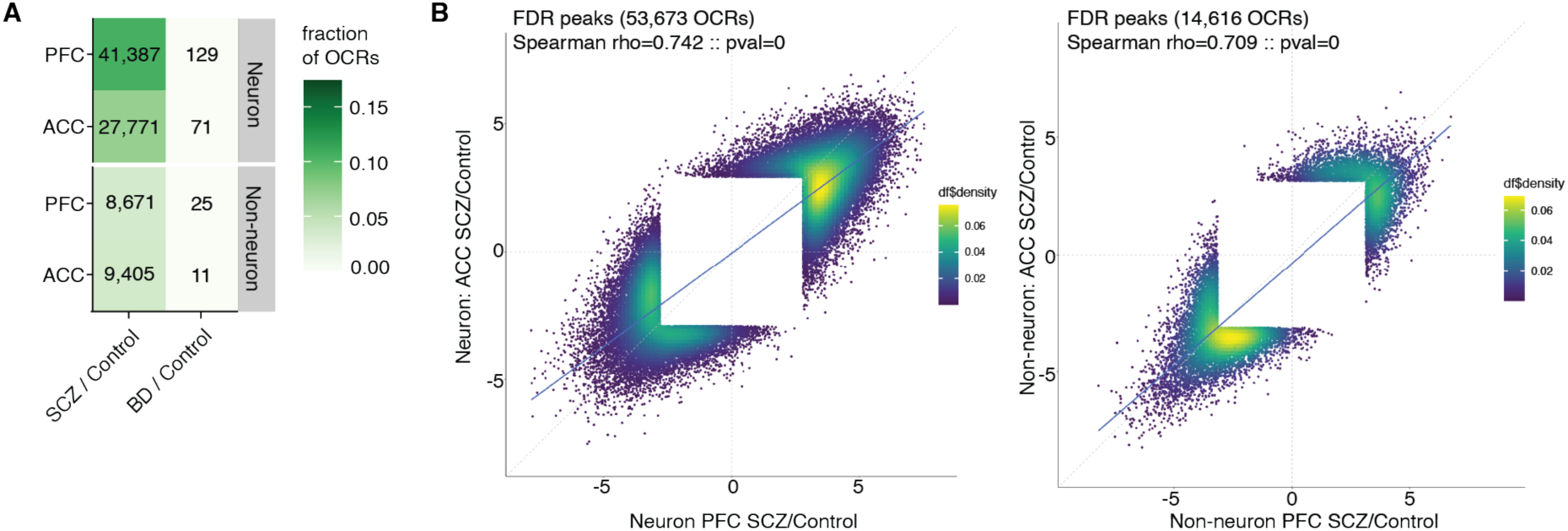
Correlation of SCZ and BD specific OCRs. **A)** Heatmap of the number of differential OCRs at FDR < .05 in for all 4 datasets: neuron PFC, neuron ACC, non-neuron PFC and non-neuron ACC across SCZ/controls and BD/controls associated. **B)** Comparison of t-statistics of significant SCZ associated changes at FDR < .05 across cortical regions in neuronal (53,673 OCRs) and non-neuronal (14,616 OCRs). White square near the origin corresponds to the OCRs that did not pass the FDR cutoff.

**Figure S10.**
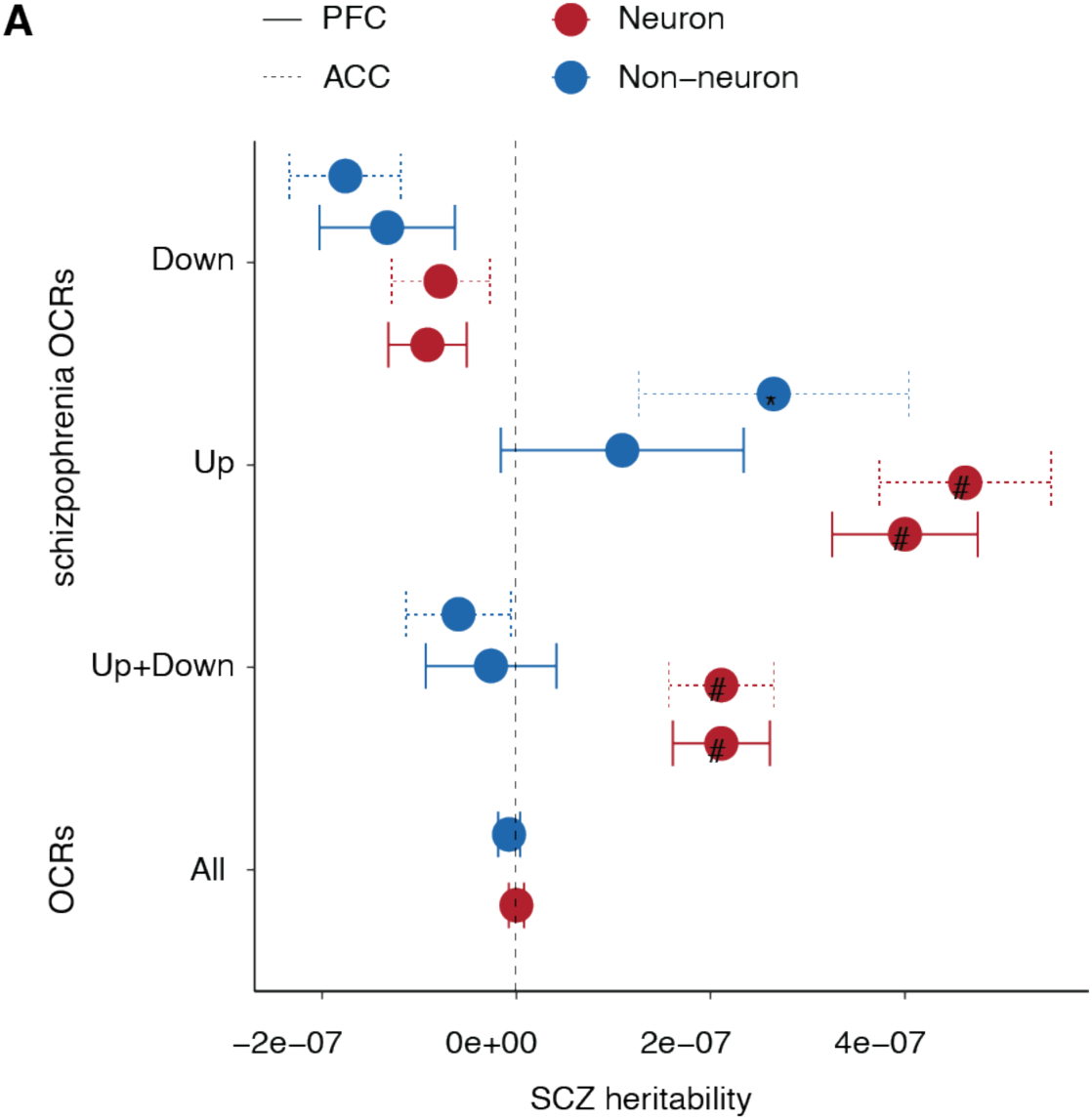
SCZ heritability of cell and region specific schizophrenia OCRs. **A**) Schizophrenia heritability coefficients of common risk variants that overlap with (1) ‘All OCRs’: all identified OCRs (391,420 in neurons (red) and 260,431 in non-neurons (blue)), (2) “Up+Down”: upregulated and downregulated schizophrenia OCRs (41,387(27,771) in PFC(ACC) neurons (red), 8,671(9,405) in PFC(ACC) non-neurons (blue)) (3) “Up”: upregulated schizophrenia OCRs (25,146(15,791) in PFC(ACC) in neurons (red), 3,726(3,075) in PFC(ACC) non-neurons (blue)); and (3) “Down”: dysregulated schizophrenia OCRs (16,241(11,980) in PFC(ACC) neurons (red), 4,945(6,330) in PFC(ACC) non-neurons (blue)). All upregulated and downregulated OCRs are with log_2_FC (schizophrenia versus controls) >0 and <0, respectively. Error bars represent standard error in schizophrenia heritability from LD score regression.

**Figure S11.**
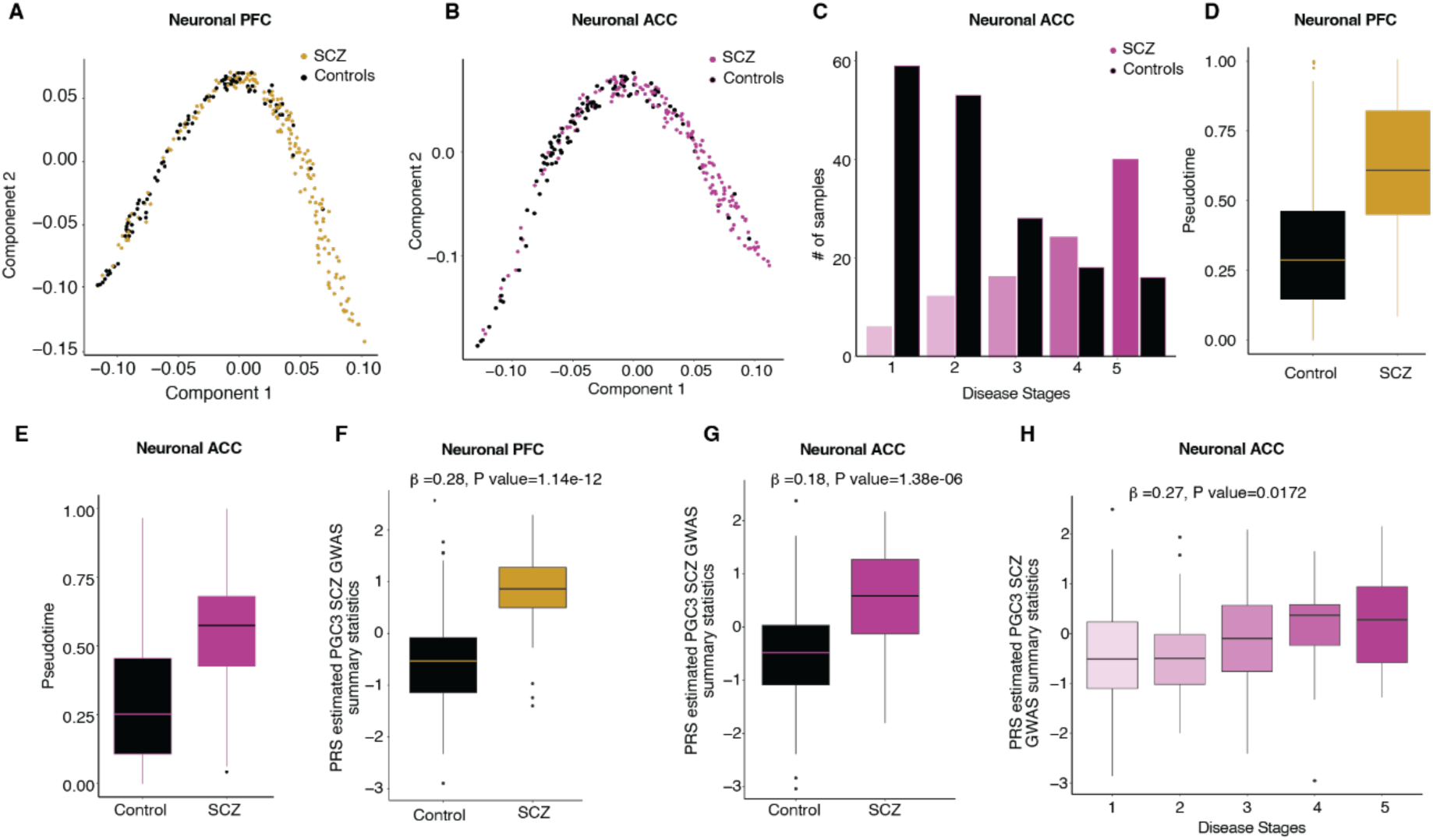
Inferred SCZ stages from chromatin accessibility generated from PFC and ACC in neurons. A-B) Estimated low-dimensional space to order samples based on similarity in expression of upregulated neuronal OCRs (n=2,411(1,495)) in SCZ from PFC(ACC) regions. The samples are colored by clinical diagnosis. SCZ samples are in yellow(purple) and controls are in black(black) in PFC(ACC) regions. **C)** Stratification of samples by clinical diagnosis as a function of inferred disease stage, where early stage (n=1) to late stage (n=5) is from left to right. **D-E)** Distribution of pseudotime for neuronal PFC and neuronal ACC samples in C and D respectively. Logistic regression model: schizophrenia/Controls ∼ pseudotime; OR=2.9, P value = 1.69e-05 in PFC (D) and OR=2.8, P value = 9.50e-05 in ACC (E). **F**-**G)** Plot of PRS of samples calculated using PGC3 SCZ GWAS summary statistics, stratified by inferred disease status in PFC and ACC region respectively. Beta estimate and p value in G-H are obtained using linear regression model: disease stages/status ∼ PRS + Age + Age^2^ + Sex. **H)** Plot of PRS of samples calculated using PGC3 SCZ GWAS summary statistics, stratified by inferred disease severity stages in ACC region. Box plots in D-H have lower and upper hinges at the 25th and 75th percentiles and whiskers extending to, at most, 1.5xIQR (interquartile range).

**Figure S12.**
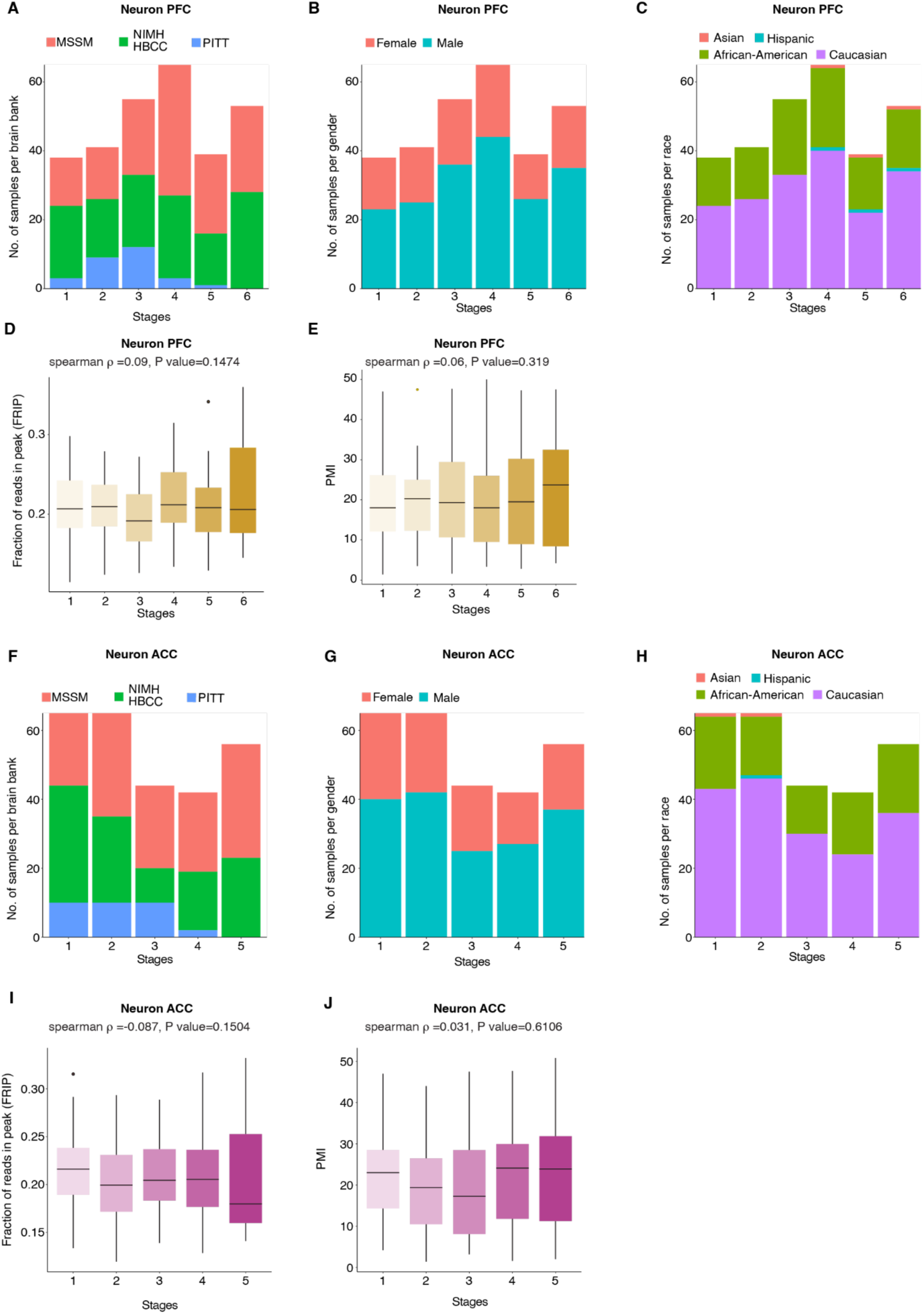
Distribution of technical and clinical variables of samples stratified by inferred disease stages. **A-C)** Bar plot to show the brain-bank, gender and race of neuron PFC samples stratified by inferred disease stages. **D-E)** Distribution of fraction of reads in peak(frip) and postmortem interval (PMI) in hrs of neuron PFC samples as a function of inferred disease stages. **F-H)** Bar plot to show the brain-bank, gender and race of neuron ACC samples stratified by inferred disease stages. **I-J)** Distribution of fraction of reads in peak (frip) and postmortem interval (PMI) in hrs of neuron ACC samples as a function of inferred disease stages. Box plots in D-E) and I-J) have lower and upper hinges at the 25th and 75th percentiles and whiskers extending to, at most, 1.5xIQR (interquartile range).

**Figure S13.**
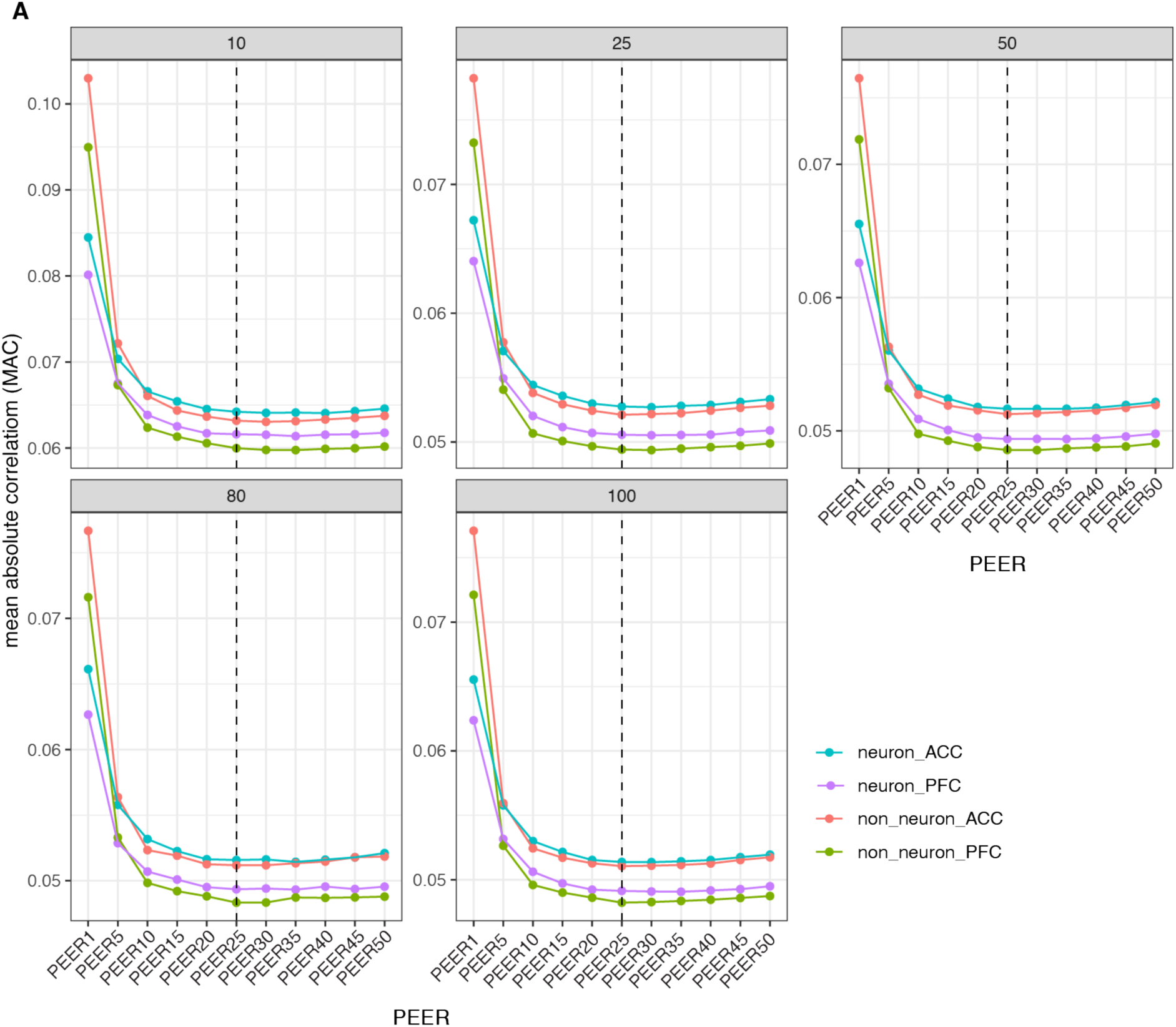
Optimal number of PEER to call final CRDs. **A)** Plot of mean absolute correlation (MAC) cut off value which is the value at 95th percentile from MAC distribution of MAC of each CRDs and permuted CRDs at i^th^ PEER.

**Figure S14.**
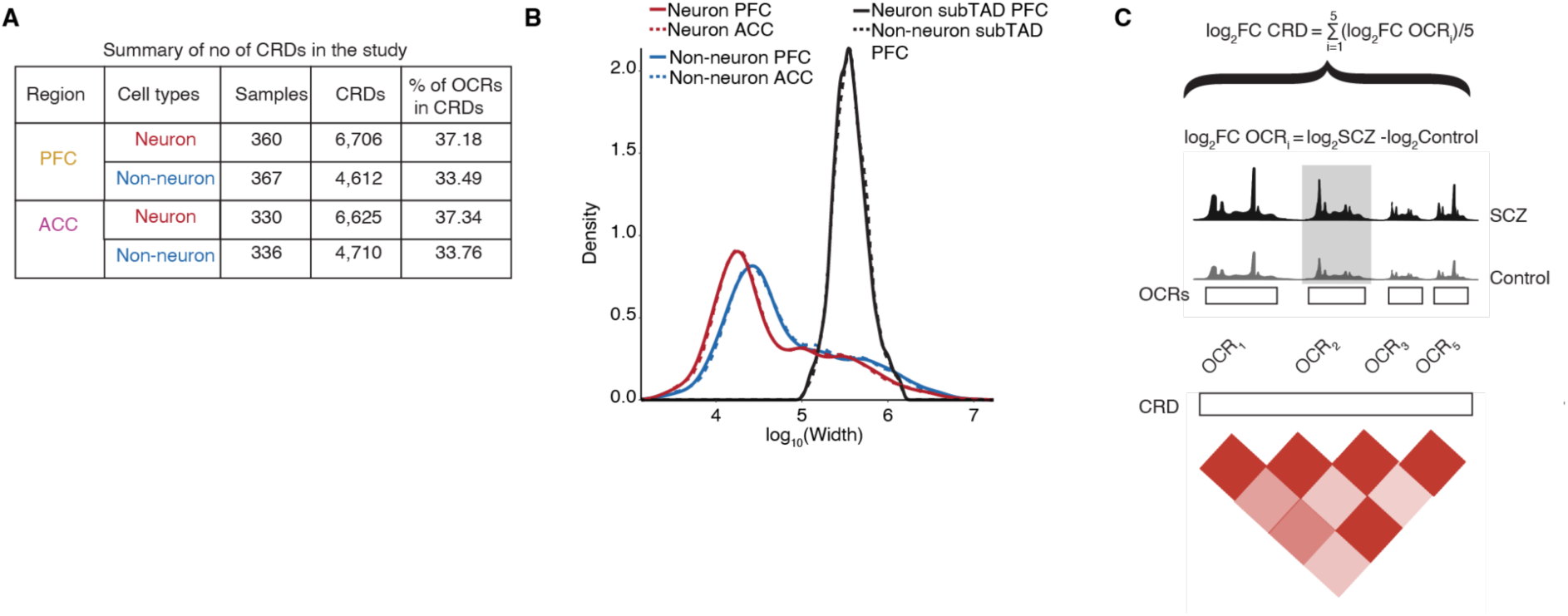
*Cis*-regulatory landscape defined by CRDs. **A**) Summary table of the number of samples, OCRs and CRDs identified across four datasets 1) neuronal PFC, 2) neuronal ACC, 3) non-neuronal PFC and 4) non-neuronal ACC. **B**) Genome-wide base pair length distribution (x-axis log scale) of CRDs (colored curves) compared to neuronal and non-neuronal PFC subTADs (black curve). **D)** Schematic of a CRD obtained from the pairwise correlation of four OCRs. The log_2_FC CRD is summarized as the average of log_2_FC (SCZ versus controls) of four OCRs.

**Figure S15.**
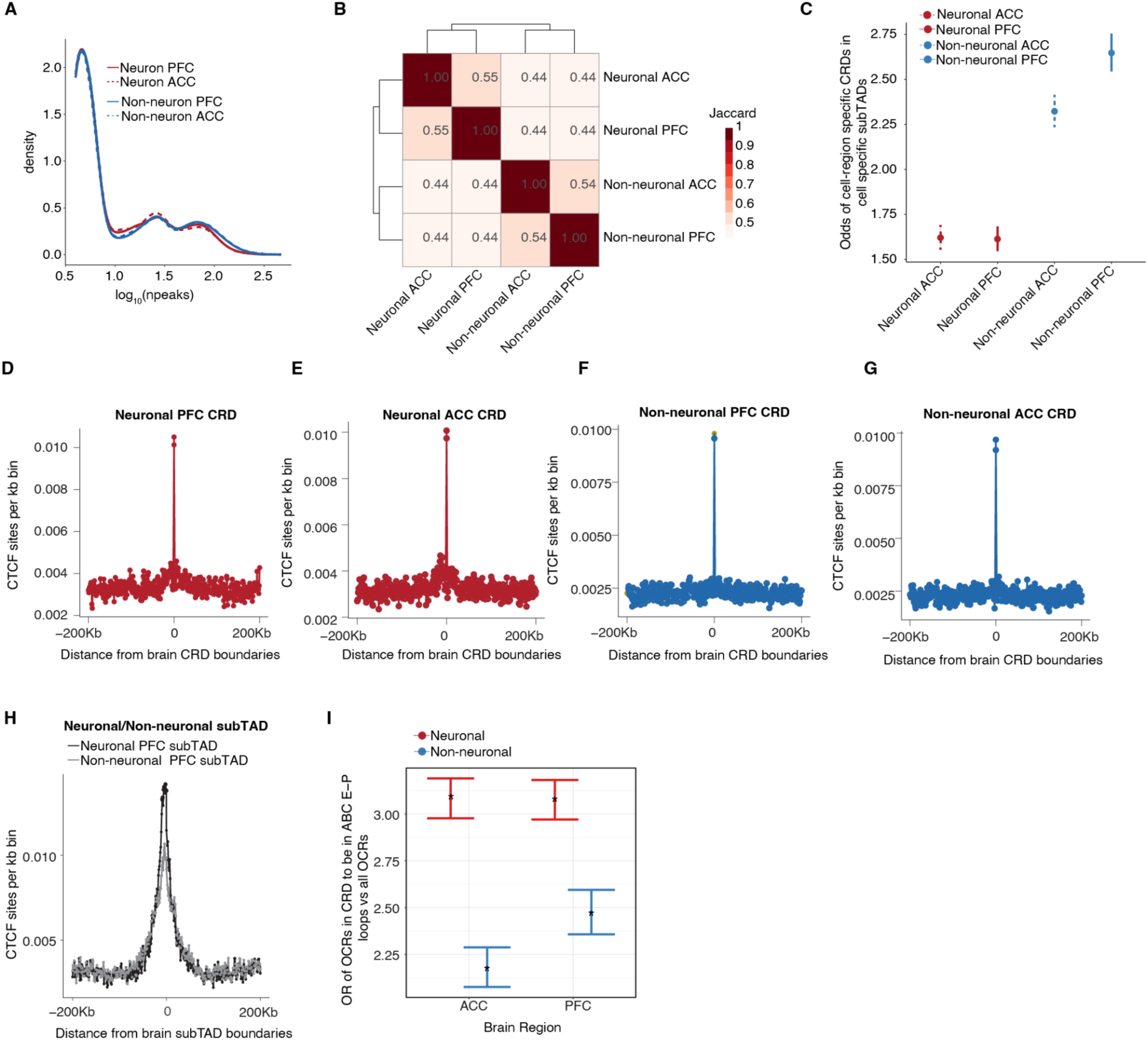
Higher order chromatin structure related characteristics of CRDs. **A**) Distribution of number of OCRs per CRD in neuron PFC, neuron ACC, non-neuron PFC and non-neuron ACC CRDs. The x-axis is in log scale. **B)** Jaccard index to show the overlap of OCRs within the cell and region specific CRDs. **C)** Odds ratios to find the cell-region specific OCRs within CRDs inside PFC cell specific subTADs when compared to all OCRs. Proportion of CTCF sites in a bin of 1Kb as a function of distance from boundaries of neuronal CRD in **D)** PFC (red), **E)** ACC (red), **F)** non-neuronal CRD PFC (blue), **G)** ACC (blue), **H)** PFC neuron subTAD (black) and PFC non-neuronal subTAD(gray). CTCF regions are from H1 stem-cell-differentiated neuronal culture in relation to distance from CRD. **I)** Odds ratio to find ABC OCRs within neuronal PFC(red), neuronal ACC(red) and non-neuronal PFC(blue), non-neuronal ACC(blue) CRDs. (neurons: OR>3, Fisher’s exact test p value < 0.05) and non-neurons CRDs (OR>2, Fisher’s exact test p value < 0.05)

**Figure S16.**
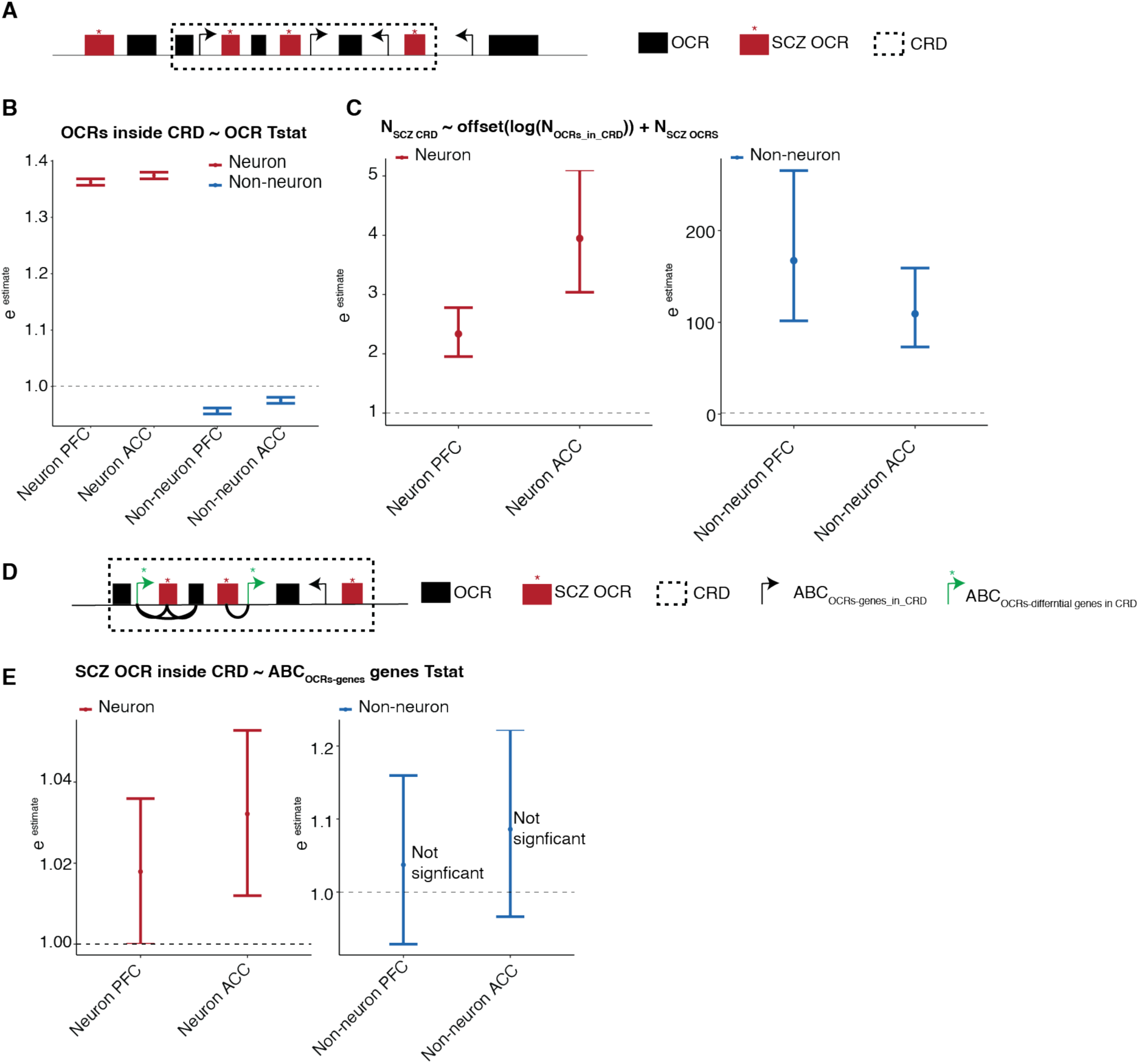
Disease relevance of CRDs. **A)** Schematic representation showing schizophrenia OCRs in red color with a “*,” while non-schizophrenia OCRs are depicted in black. An example of a CRD is enclosed within a dashed box, and black arrows indicate TSS sites of genes. **B)** Coefficient of the logistic regression-based generalized linear model used to predict the status of neuronal (non-neuronal) OCRs located inside or outside neuronal (non-neuronal) CRDs in both cortical regions colored in red(blue). This prediction is made using SCZ-associated t-statistics obtained from the cell-region specific differential OCRs analysis. **C)** Coefficient of the Poisson-based glm (generalized linear model) to predict whether a CRD is a SCZ CRD or not using the number of schizophrenia OCRs within the CRD as an independent variable. An offset term of the number of OCRs within the CRD was included in the model. **D)** Schematic representation showing schizophrenia OCRs in red with “*”, and non-schizophrenia OCRs in black. An example of a CRD and OCRs within it are enclosed within a dotted box. Black arrows show TSS sites of genes mapped to OCRs, while green arrows indicate TSS sites of ABC differential genes in SCZ vs. Controls, which are mapped to OCRs. **E)** Coefficient of the logistic regression-based generalized linear model used to predict the status of neuronal (non-neuronal) schizophrenia OCRs located inside or outside neuronal (non-neuronal) CRDs in both cortical regions colored in red(blue). SCZ-associated t-statistics obtained from the differential genes mapped to OCRs within the CRD is an independent variable in the model. The differential genes analysis is of CMC cohort(*19*).

**Figure S17.**
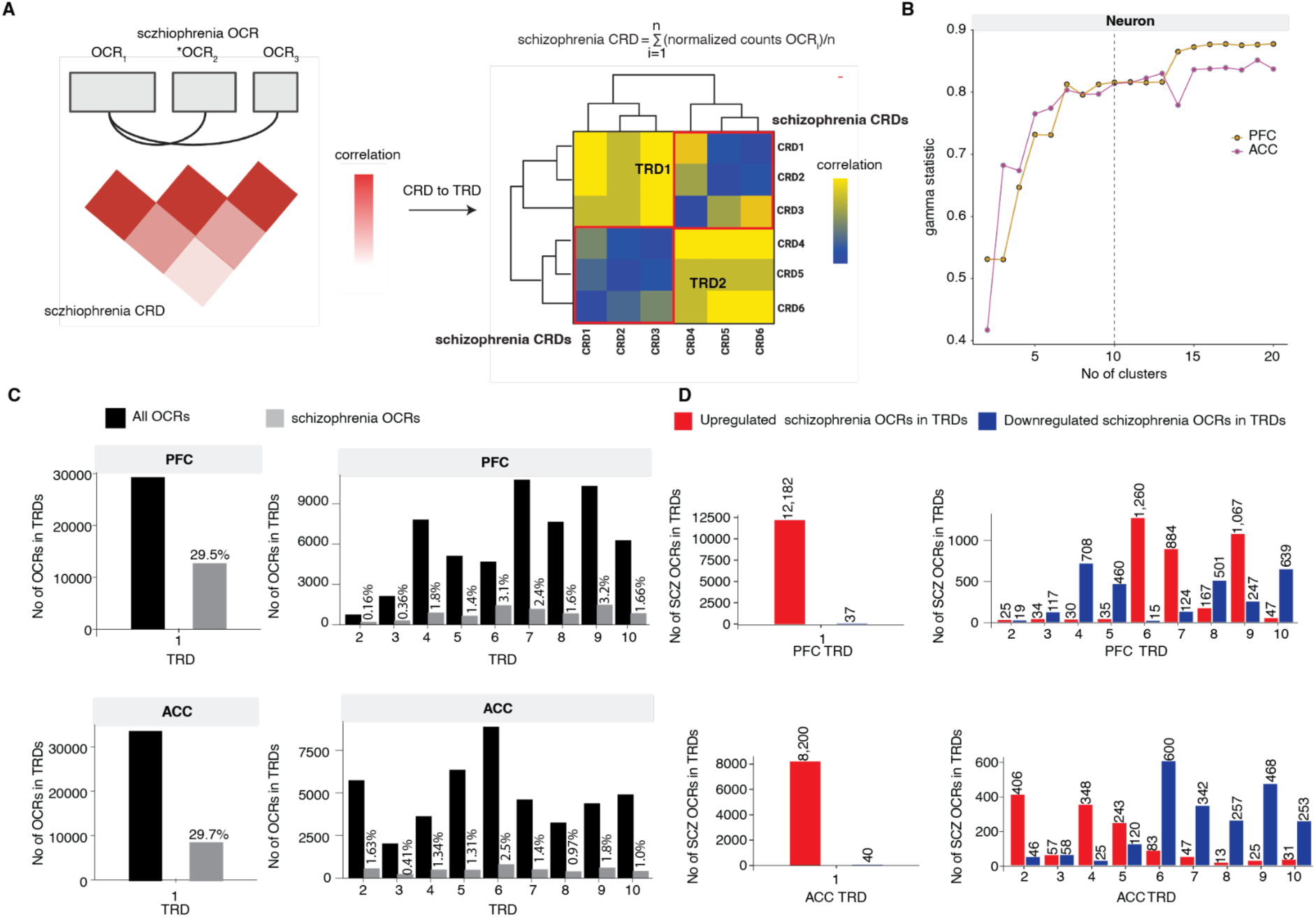
Characteristics of trans-regulatory domains of CRDs. **A)** Schematic workflow depicting identification of TRDs from hierarchical clustering of schizophrenia CRDs. **B)** Gamma statistics were plotted against the number of clusters in Chromatin Regulatory Domain (CRD) matrices of neuronal PFC and ACC. The dotted line indicates the optimal number of clusters, *n*=10, which was selected for both neuronal PFC and ACC through the hierarchical clustering method. **C)** The number of OCRs and SCZ associated OCRs are represented as “All OCRs’’ and “schizophrenia OCRs,” respectively, within CRDs in neuronal PFC TRDs (top plots) and non-neuronal ACC TRDs (bottom plots). **D)** The number of upregulated and downregulated schizophrenia OCRs within TRDs are depicted separately in neuronal PFC TRDs (top plots) and neuronal ACC TRDs (bottom plots).

**Figure S18.**
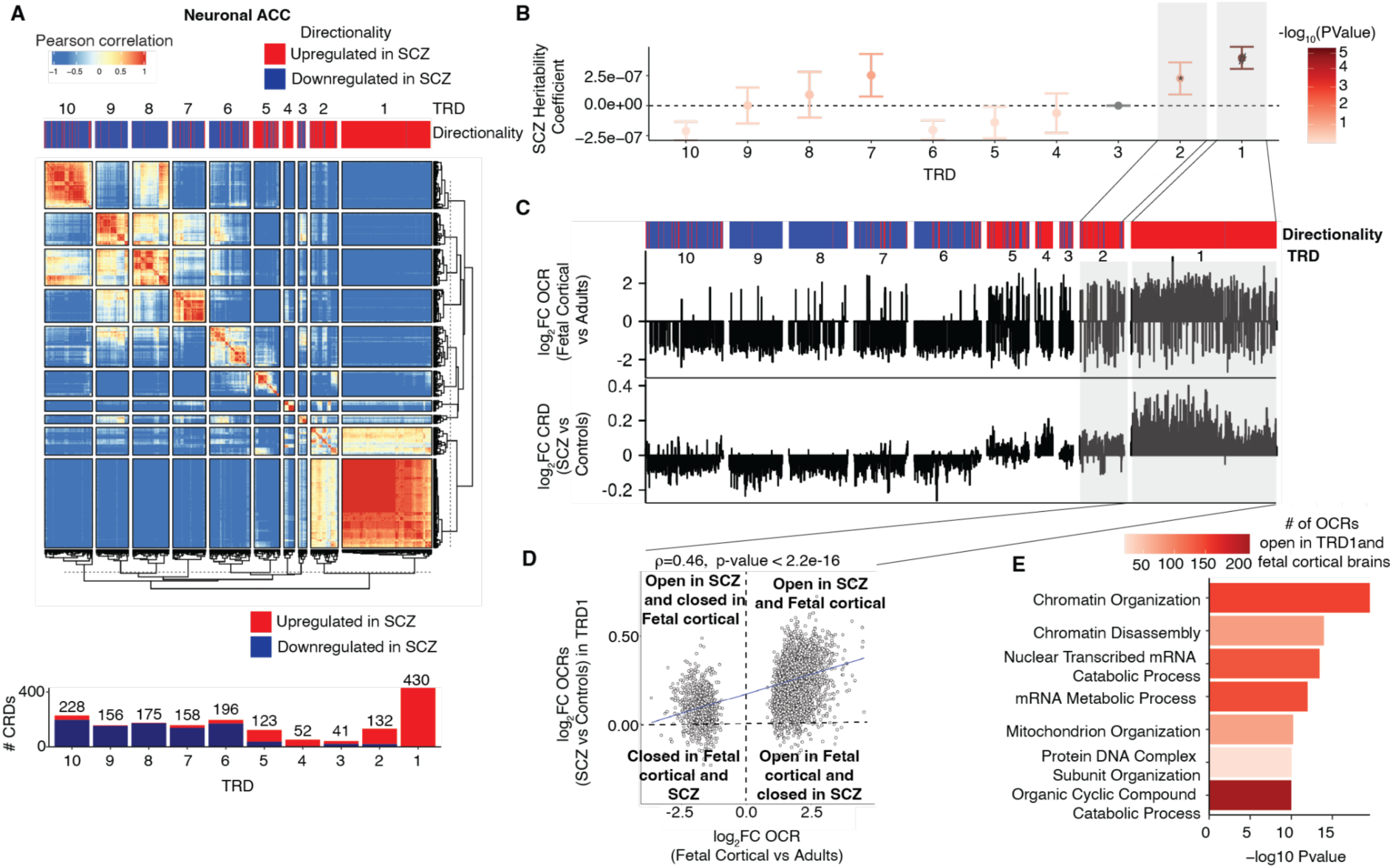
Trans regulatory domains from hierarchical clustering of neuronal SCZ CRDs from the ACC. **(A)** Heatmap depicting hierarchical clustering of trans interaction of 1,691 SCZ CRDs across 330 samples from the ACC. The clustering results in ten trans regulatory domains (TRDs) using gamma statistics. The directionality annotation bar plot above the heatmap shows upregulation in red (log_2_SCZ-log_2_Control >0) and downregulation in navy (log_2_SCZ-log_2_Control >0) of SCZ neuronal CRDs. Bottom plot shows a barplot of the number of up and downregulated SCZ neuronal CRDs per TRD from the ACC region. **(B)** Coefficients of SCZ heritability stratified by TRDs. The overlap of OCRs within the TRDs with SCZ risk variants was assessed using LD score regression. *P* values are from LD score regression. ‘#’: significant for enrichment in LD score regression after Benjamini–Hochberg FDR correction for multiple testing across all tests in the plot (FDR < 5%). Error bars show standard error in SCZ heritability from LD score regression. **(C)** Top and bottom bar plot show log_2_FC_fetal_cortical_ (Fetal cortical vs adult controls) and log_2_FC (SCZ vs Controls) of neuronal ACC CRD respectively **(D)** Spearman correlation of log_2_FC_fetal_cortical_ compared to log_2_FC (SCZ vs Controls) of neuronal ACC CRDs from TRD1 (n=3,149 OCRs). P value is obtained from Spearman’s rank correlation (ρ) test. **(E)** Functional pathway enrichment of OCRs from neuronal ACC TRD1 that overlap with fetal cortical specific OCRs.

**Figure S19.**
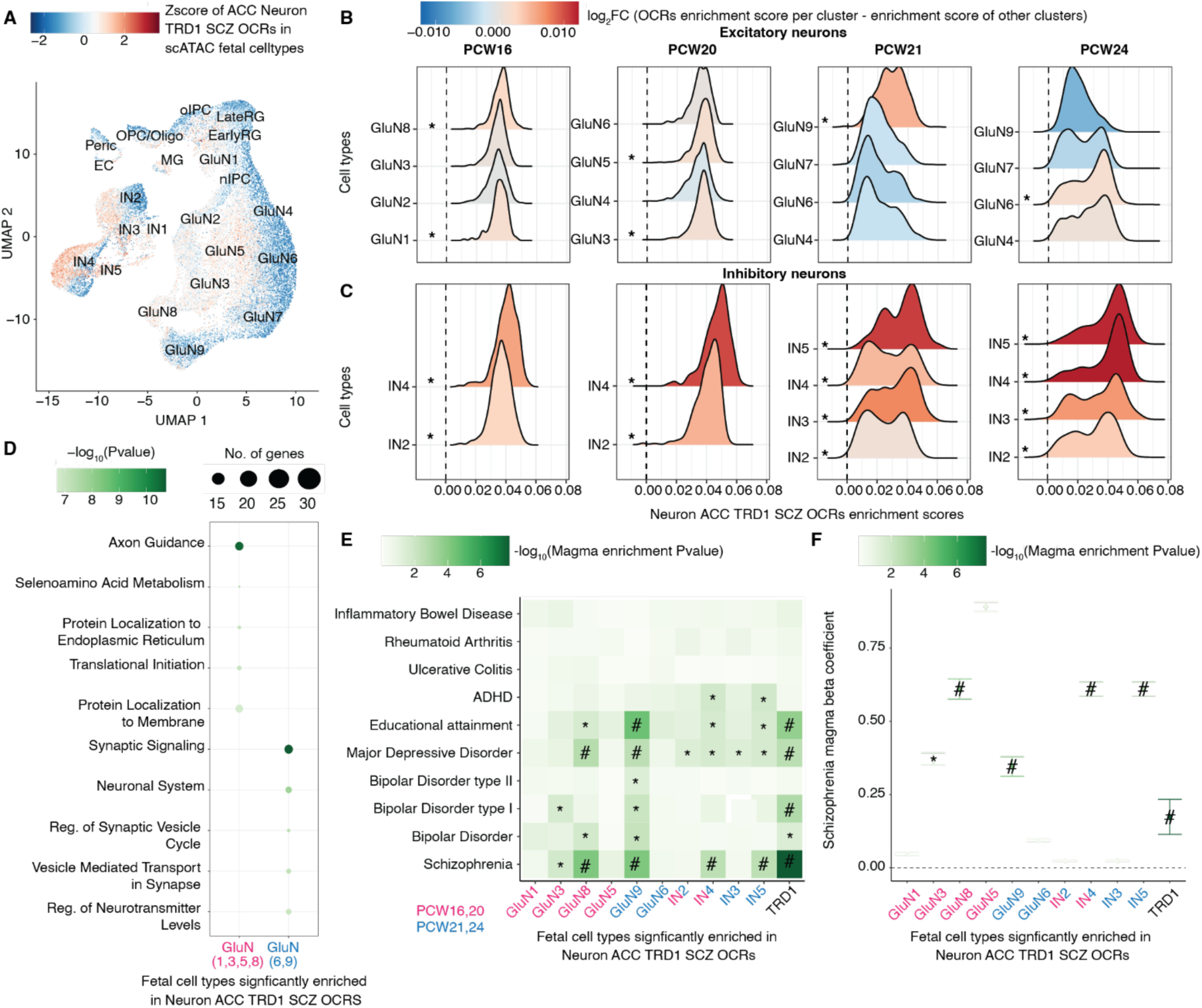
Analysis of cell-type specificity of neuronal ACC schizophrenia OCRs in fetal cortical scATAC-seq data. **A**) UMAP plot of fetal cortical scATAC-seq data from Trevino *et al.* in which each cell type is colored by Z-scores of schizophrenia OCRs in neuronal ACC TRD1. The cell types include early radial glia (EarlyRG), late radial glia (LateRG), oligodendrocyte progenitor cell/oligodendrocyte (OPC/Oligo), neuronal intermediate progenitor cell (nIPC), oligo intermediate progenitor cell (oIPC), glutamatergic neuron (GluN), interneuron (IN), endothelial cell (EC), microglia (MG), and pericytes (Peric). **B**) Distribution of enrichment scores stratified by two major classes of cell types: 1) excitatory neurons (nine glutamatergic neurons) and 2) inhibitory neurons (five inhibitory neurons). The color represents the magnitude of coefficient of association of enrichment scores of schizophrenia OCRs from neuronal ACC TRD1 at each developmental stage separately for each cell types, using the model (enrichment score ∼ celltype + (1| sample ID) where cell types are grouped as the cell type of interest vs. all other cell types. * depicts the cell types that are significantly enriched from the latter test and also have higher enrichment than the schizophrenia OCRs outside TRDs. **C**) Functional pathway analysis conducted on the early and late fetal glutamatergic specific marker genes that are also annotated by schizophrenia OCRs in TRD1. **D**) Heat map of MAGMA enrichment *P* values of brain-related and non-brain related GWAS traits and **E**) Coefficients of enrichment of common SCZ risk variants using MAGMA in fetal cell specific marker genes that are also annotated to schizophrenia OCRs within neuronal ACC TRD1. The x-axes in **D)** and **E)** are 1) early fetal (16+20 pcw) glutamatergic (GluN 1,3,5,8) in magenta, 2) late fetal (21+24 pcw) glutamatergic (GluN 6,9) in blue, 3) early fetal (16+20 pcw) inhibitory (IN2,4) in magenta, 4) late fetal (21+24 pcw) inhibitory (IN 2,3,4,5) in blue fetal cell types and all ABC mapped schizophrenia OCRs to genes within neuronal ACC TRD1 in black. ‘#’: significant MAGMA enrichment coefficient after FDR correction of multiple testing across all tests in the plot (Benjamini–Hochberg test); ‘*’: nominally significant for enrichment in (**D**) and (**E**).

**Figure S20.**
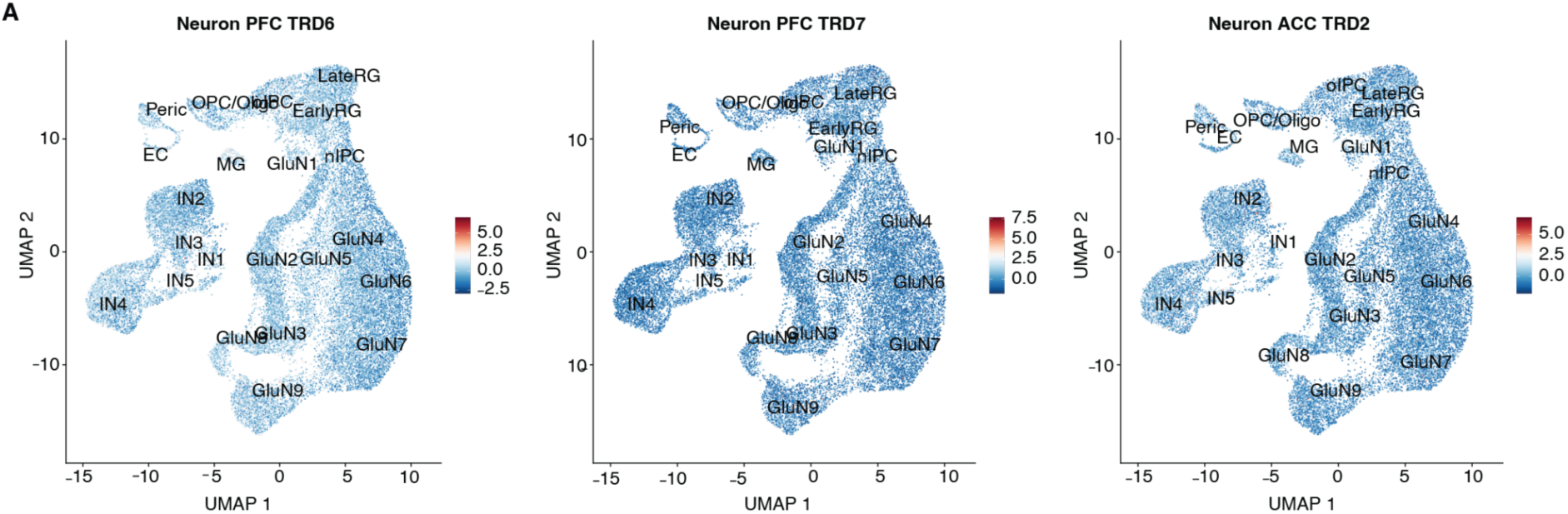
Enrichment of expression of schizophrenia OCRs in SCZ TRDs in fetal cortical OCRs in scATAC-seq data. **A**) UMAP plot of fetal cortical scATAC-seq data from Trevino *et al.* in which each cell type is colored by Z-scores of neuronal PFC schizophrenia OCRs in neuronal PFC TRD6, neuronal PFC TRD7 and neuronal ACC schizophrenia OCRs in neuronal ACC TRD2. The cell types include early radial glia (EarlyRG), late radial glia (LateRG), oligodendrocyte progenitor cell/oligodendrocyte (OPC/Oligo), neuronal intermediate progenitor cell (nIPC), oligo intermediate progenitor cell (oIPC), glutamatergic neuron (GluN), interneuron (IN), endothelial cell (EC), microglia (MG), and pericytes (Peric).

**Figure S21.**
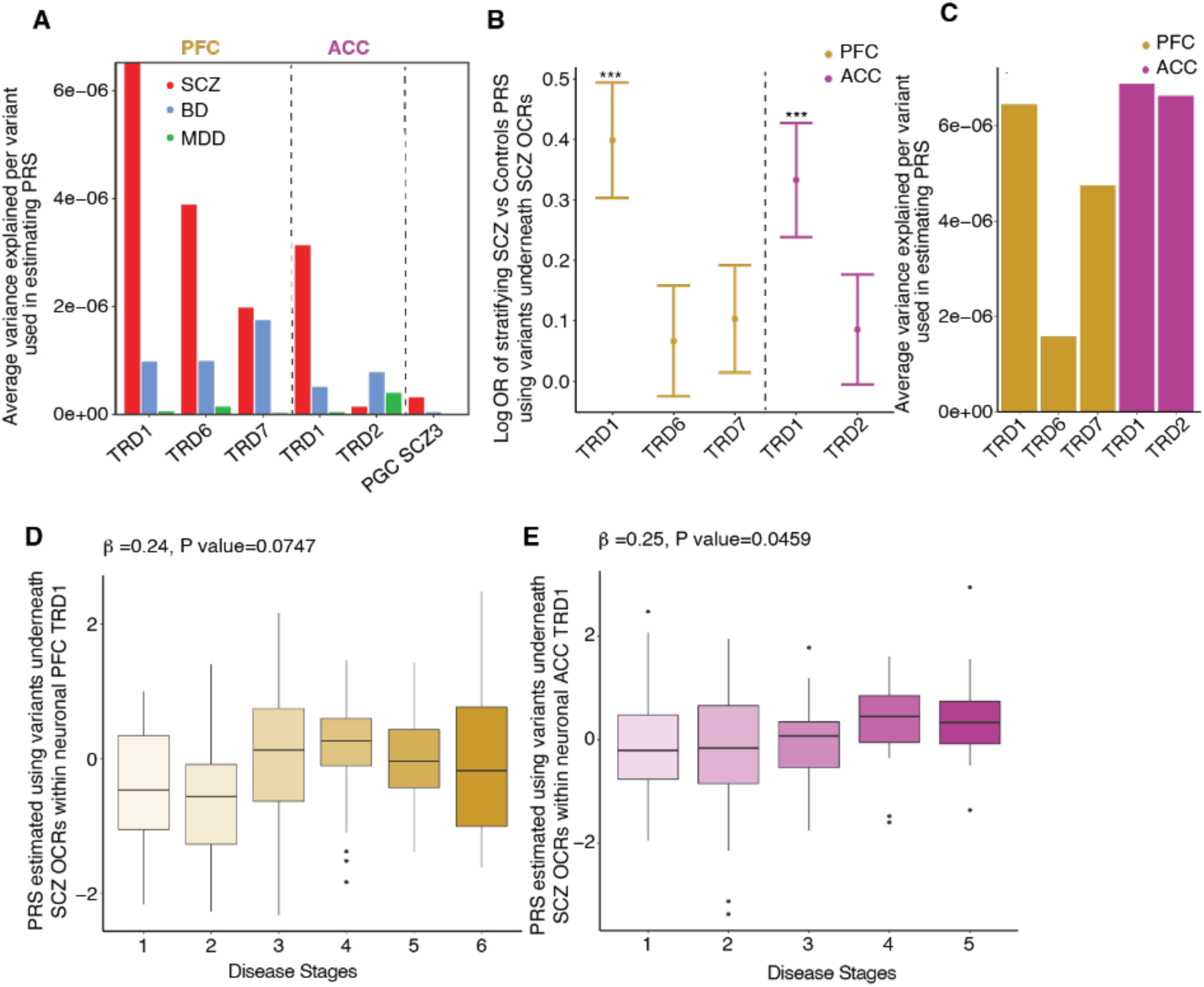
Trans Regulatory Domain one (TRD1) in PFC and ACC can stratify SCZ cases and controls in MVP and CMC cohort. For SCZ TRDs: TRD1, TRD6 and TRD7 from PFC region and TRD1, TRD2 from ACC region. **A**) Average variance explained per variant used in estimating PRS of MVP cohort (PFC TRDs: 1,388 for TRD1, 139 for TRD6 and 103 for TRD7; ACC TRDs: 815 for TRD1 and 46 for TRD2; PGC3 SCZ: 350,882). **B**) Plot of logOR from logistic regression of PRS of 632 individuals from CMC cohort estimated using the SNPs in OCRs from these TRDs to predict their case and control status, **C**) Average variance explained per variant used in estimating PRS of CMC cohort (PFC TRDs: 3,869 for TRD1, 449 for TRD6 and 357 for TRD7; ACC TRDs: 2,447 for TRD1 and 172 for TRD2; PGC3 SCZ: 350,882). **D-E)** Turkey bar plot of PRS of samples calculated using the SNPs underneath schizophrenia OCRs within TRD1 (subsetted from the full PGC3 SCZ GWAS summary statistics), stratified by inferred disease severity stages in PFC and ACC respectively.

### Supplementary Tables

**Table S1:** Metadata and summary of QC metrics for neuronal PFC, neuronal ACC, non-neuronal PFC and non-neuronal ACC samples.

**Table S2:** GWAS enrichment of brain traits and non-brain related traits in neuronal and non-neuronal OCRs.

**Table S3:** Inferred stages of SCZ/controls of neuronal and non-neuronal samples in PFC and ACC regions using manifold learning method and their respective polygenic risk scores estimated using PGC3 SCZ GWAS summary statistics.

**Table S4:** List of genomic coordinates of identified neuronal PFC, neuronal ACC, non-neuronal PFC and non-neuronal ACC CRDs.

**Table S5:** GWAS enrichment of brain traits and non-brain related traits in OCRs within neuron PFC, neuron ACC, non_neuron PFC and non_neuron ACC CRDs stratified by OCR location (inside or outside CRDs).

**Table S6:** GWAS enrichment of brain traits and non-brain related traits of schizophrenia OCRs that reside inside and outside of neuronal PFC, neuronal ACC, non-neuronal PFC and non-neuronal ACC CRDs.

**Table S7:** Association test of scFetal cell types with neuronal TRD expression scores in PFC and ACC regions. Magma enrichment of brain and non-brain related traits in significantly associated scfetal cell types from association test with TRD1 marker genes that are present within the neuron TRD1 in PFC and ACC regions. List of TRD1 associated scfetal cell types marker genes and their functional pathways analysis using GREAT tool.

